# Investigation of choroid plexus variability in schizophrenia-spectrum disorders – insights from a multimodal study

**DOI:** 10.1101/2023.12.18.23300130

**Authors:** Vladislav Yakimov, Joanna Moussiopoulou, Lukas Roell, Marcel S. Kallweit, Emanuel Boudriot, Matin Mortazavi, Sergi Papiol, Lenka Krčmář, Mattia Campana, Eva C. Schulte, Nicolas Glaichenhaus, Emanuela Martinuzzi, Sean Halstead, Nicola Warren, Dan Siskind, Isabel Maurus, Alkomiet Hasan, Peter Falkai, Andrea Schmitt, Florian Raabe, CDP Working Group, Daniel Keeser, Elias Wagner

**Author notes:** **Corresponding author:** Dr. med. Vladislav Yakimov, Address: Department of Psychiatry and Psychotherapy, University Hospital, LMU Munich, Nussbaumstrasse 7, 80336 Munich, Germany. CDP Working Group: Valéria de Almeida, Stephanie Behrens, Emanuel Boudriot, Mattia Campana, Fanny Dengl, Peter Falkai, Laura E. Fischer, Nadja Gabellini, Vanessa Gabriel, Thomas Geyer, Katharina Hanken, Alkomiet Hasan, Genc Hasanaj, Alexandra Hirsch, Georgios Ioannou, Iris Jäger, Sylvia de Jonge, Marcel S. Kallweit, Temmuz Karali, Susanne Karch, Berkhan Karslı, Daniel Keeser, Christoph Kern, Nicole Klimas, Maxim Korman, Lenka Krčmář, Isabel Lutz, Julian Mechler, Verena Meisinger, Matin Mortazavi, Joanna Moussiopoulou, Karin Neumeier, Frank Padberg, Boris Papazov, Sergi Papiol, Pauline Pingen, Oliver Pogarell, Siegfried Priglinger, Florian J. Raabe, Lukas Roell, Moritz J. Rossner, Andrea Schmitt, Susanne Schmölz, Enrico Schulz, Benedikt Schworm, Elias Wagner, Sven Wichert, Vladislav Yakimov, Peter Zill. These authors contributed equally (Daniel Keeser, Elias Wagner).

## Abstract

**Background and Hypothesis:** Previous studies have suggested that choroid plexus (ChP) enlargement occurs in individuals with schizophrenia-spectrum disorders (SSD) and is associated with peripheral inflammation. However, it is unclear whether such an enlargement delineates a biologically defined subgroup of SSD. Moreover, it remains elusive how ChP is linked to brain regions, associated with peripheral inflammation in SSD.

**Study Design:** A cross-sectional cohort of 132 individuals with SSD and 107 age-matched healthy controls (HC) underwent magnetic resonance imaging (MRI) of the brain and clinical phenotyping to investigate the ChP and associated regions. Case-control comparison of ChP volumes was conducted and structural variance was analysed by employing the variability ratio (VR). K-means clustering analysis was used to identify subgroups with distinct patterns of the ventricular system and the clusters were compared in terms of demographic, clinical and immunological measures. The relationship between ChP volumes and brain regions, previously associated with peripheral inflammation, was investigated.

**Study Results:** We could not find a significant enlargement of the ChP in SSD compared to HC but detected an increased VR of ChP and lateral ventricle volumes. Based on these regions we identified 3 clusters with differences in age, symbol coding test scores and possibly inflammatory markers. Larger ChP volume was associated with higher volumes of hippocampus, putamen, and thalamus in SSD, but not in HC.

**Conclusions:** This study suggests that ChP variability, but not mean volume, is increased in individuals with SSD, compared to HC. Larger ChP volumes in SSD were associated with higher volumes of regions, previously associated with peripheral inflammation.

## 1. Introduction

Schizophrenia-spectrum disorders (SSD) are severe mental illnesses with heterogenous clinical presentation and pathophysiology, as indicated by increased brain structural variability^1, 2^. This heterogeneity has largely contributed to the limited mechanistic understanding of SSD and has arguably led to a decrease in novel drug development efforts by pharmaceutical companies^3^. There is therefore an unmet need for the identification of *intermediate phenotypes*, which can help delineate subgroups within the schizophrenia-spectrum with common pathophysiological mechanisms. Such *intermediate phenotypes* could aid gaining mechanistic understanding in defined SSD subgroups and pave the way to a more personalised treatment approach^4^.

Recent studies indicate that dysregulation of the immune system is a contributing factor in a subgroup of people with SSD^5, 6^. It may affect brain structure and functioning, potentially resulting in increased risk for more pronounced symptom severity^7^. Furthermore, increased levels of peripheral inflammatory markers have been consistently described in SSD^7, 8^. Some of those alterations have been associated with increased blood-cerebrospinal fluid (CSF) barrier (BCB) permeability^9^, which is a common finding in SSD^10–14^ and suggests concomitant neuroinflammation^10, 15^. Nevertheless, there is substantial inconsistency regarding the number of proposed inflammatory subgroups in SSD and their impact on brain structure, with some studies indicating increases, while others show reductions in regional (sub-)cortical volumes^16,17^. Recent data suggests the presence of five distinct inflammatory subgroups of chronic schizophrenia, with one cluster characterized by elevated levels of IFN-γ. This cluster is associated with attenuated brain atrophy and cognitive dysfunction, suggesting protective properties of some types of inflammation^18^.

In line with these findings, increased ChP volumes, altered ChP epithelia, and associated upregulation of immune genes in the ChP were described in individuals with SSD^19–21^. Located in each brain ventricle, the ChP produces CSF and forms the BCB, thus regulating central nervous system (CNS) homeostasis^22^. In recent years, its role as a unique neuroimmunological niche has been increasingly recognized^23^. Its ability to sense and engage signals from the periphery and CNS parenchyma and to host different immune cell populations stresses its role as a central hub of the brain-body interface^23–25^. ChP epithelial cells interact with peripheral leukocytes and can release cytokine effectors in the context of acute inflammation or attenuate the immune response in chronic inflammation, thus modulating peripheral and CNS immunity^24^. ChP can also exert neuroprotective properties via the secretion of neurotrophic factors ^26^.

Increased ChP volume has been consistently demonstrated in neurodegenerative and neuroinflammatory disorders and has been linked to brain atrophy, neurodegeneration, lesion expansion and disability progression^27–29^. A recent study indicated a volumetric increase of the ChP in people across the psychosis spectrum and linked it to worse cognition, brain atrophy and peripheral inflammation^19^. Moreover some preliminary findings suggest that the ChP volume increase was present in early, but not in chronic psychosis^30^. Due to the preliminary nature of those findings, they require further confirmation. Moreover, there is a substantial knowledge gap in the role of the ChP and the associated ventricular system in SSD. Most of the previous studies focused on mean volumetric differences of the ChP between individuals with psychotic disorders and healthy controls (HC) but did not explore its structural variability. This is especially important in light of the mounting evidence for inflammatory subgroups in SSD and the strong implication of ChP in neuroinflammatory processes. Furthermore, no study so far has explored the relationship between ChP volume and brain structures, associated with peripheral inflammation in SSD^16^.

In the context of the described evidence gaps, we aimed to 1) replicate the findings of enlarged ChP in SSD, 2) explore the structural variance of ChP and associated ventricular regions in SSD, 3) identify distinct clusters based on the regions with increased variance and characterize them, and 4) investigate the relationship between ChP volume and volumes of subcortical and cortical regions, previously associated with peripheral inflammation in SSD.

## 2. Methods

### 2.1 Participants

This project was carried out in the context of the Clinical Deep Phenotyping (CDP) study^31^, an ongoing add-on study to the Munich Mental Health Biobank (project number 18-716)^32^ (German Clinical Trials Register, DRKS, ID: DRKS00024177) and approved by the ethics committee of the Faculty of Medicine, LMU University Hospital Munich (project numbers 20-0528 and 22-0035).

Study participants were recruited at the Department of Psychiatry and Psychotherapy, LMU University Hospital, LMU Munich, Germany, between July 14, 2021, and May 22, 2023. In- (N=76) and outpatients (N=56) were included. Healthy controls were recruited from the local community via announcements in different channels (e.g., homepage of the university hospital, local advertising). All study participants were between 18 and 65 years old and provided written informed consent. Mini International Neuropsychiatric Interview (M.I.N.I.)^33^, German version 7.0.2, based on DSM-5 criteria, was performed with all study participants as previously described^34^. Included patients had a primary diagnosis of schizophrenia, schizoaffective disorder, delusional disorder, or brief psychotic disorder, collectively referred to as SSD throughout the manuscript. Participants with no current or past psychiatric disorder according to the M.I.N.I. were defined as healthy controls (HC). HC were not screened for family history of SSD. All inpatients (76 subjects) underwent a basic blood test as part of routine clinical care. The full blood analysis included a complete blood count among others. Serum from a subgroup of individuals with SSD (78 subjects), who agreed to blood sampling, was analyzed for high-sensitivity C-reactive protein (hsCRP) with the immunoassay V-Plex Human CRP Kit (Mesoscale Discovery Kit, Catalog Nr. K151STD-1). Whole blood and serum were collected during the morning hours (8 am to 11 am), but not under fasting conditions.

Exclusion criteria were as follows: concurrent clinically relevant central nervous system (CNS) disorders, such as multiple sclerosis and epilepsy, history of encephalitis, meningitis, stroke, traumatic brain injury or cerebral surgery; current pregnancy or lactation; rheumatic disorders, inflammatory bowel disease, active malignancy, acute or chronic infection.

### 2.2. Clinical assessment

The clinical assessment was carried out as previously described by our working group^34^. Severity of psychotic symptoms was assessed with the Positive and Negative Syndrome Scale (PANSS) and global function with the Global Assessment of Functioning (GAF) scale, performed by trained raters^35^. Information regarding medication, duration of illness (DUI), duration of untreated psychosis (DUP), body mass index (BMI), concomitant somatic conditions and current smoking status was collected based on self-report and by examining medical reports. Current antipsychotic medication was translated in chlorpromazine equivalent doses (CPZeq) according to the Defined Daily Dose method^36^. A history of clozapine use was used as a proxy for treatment-resistant schizophrenia (TRS)^37^.

Cognitive performance was assessed using the German version of the Brief Assessment of Cognition in Schizophrenia (BACS)^38^. The battery includes seven cognitive tasks covering the domains of verbal memory, verbal fluency, attention and speed of information processing, working memory, motor speed, and executive function^38^. Z-scores were calculated from the raw scores of each subtest based on the means and standard deviations of our HC group. A composite z-score as a measure of global cognition was then obtained by averaging the individual z-scores of all tasks and calculating a new z-score for the average^38^. Both the subtest z-scores and composite z-score were used for statistical analyses. The BACS was performed only in participants with a native-speaker level of German.

### 2.3 Magnetic resonance imaging

All participants underwent brain magnetic resonance imaging (MRI). Brain MRI recordings were performed on a 3T Siemens Magnetom Prisma scanner (Siemens Healthineers AG, Erlangen, Germany) with a 32-channel head coil. T1-weighted scans were acquired using a magnetization-prepared rapid gradient echo (MP-RAGE) sequence with an isotropic voxel size of 0.8 mm^3^, 208 slices, repetition time of 2500 ms, echo time of 2.22 ms, flip angle of 8°, and a field of view of 256 mm. The measures of regional brain volumes (incl. choroid plexus) as well as cortical thickness were computed in mm^3^ using FreeSurfer (v7.3.2; https://surfer.nmr.mgh.harvard.edu/)^39^ and were corrected for intracranial volume^40^. The FreeSurfer atlas was used to obtain the regional volumes for left and right brain hemispheres in the following regions of interest: choroid plexus (Figure S1), lateral ventricle (sum of lateral ventricle and inferior lateral ventricle), third ventricle, fourth ventricle, hippocampus, amygdala, putamen, thalamus, anterior cingulate cortex, posterior cingulate cortex, insula, cerebellum cortex, anterior corpus callosum, fusiform gyrus, superior temporal gyrus, superior frontal gyrus, precentral gyrus, caudal middle frontal gyrus, medial orbitofrontal cortex, inferior temporal gyrus, posterior parietal cortex, lateral occipital cortex and brainstem. Those regions were selected either based on their anatomical and functional association with the choroid plexus (main regions of interest), due to their strong implication in convergent transdiagnostic and/or schizophrenia-specific brain networks (“reference regions”), as demonstrated by high-quality meta-analytic evidence^41, 42^ (Figure 1C), or due to their previous association with peripheral inflammation in schizophrenia^16^ (Table S4). The sum of the computed measures for left and right brain hemispheres of each region of interest was used for statistical analyses. The choroid plexus volume measures for comparison between SSD and HC and the variability ratios of regions of interest were additionally computed unilaterally (Table S1, S2b).

**Figure 1.**
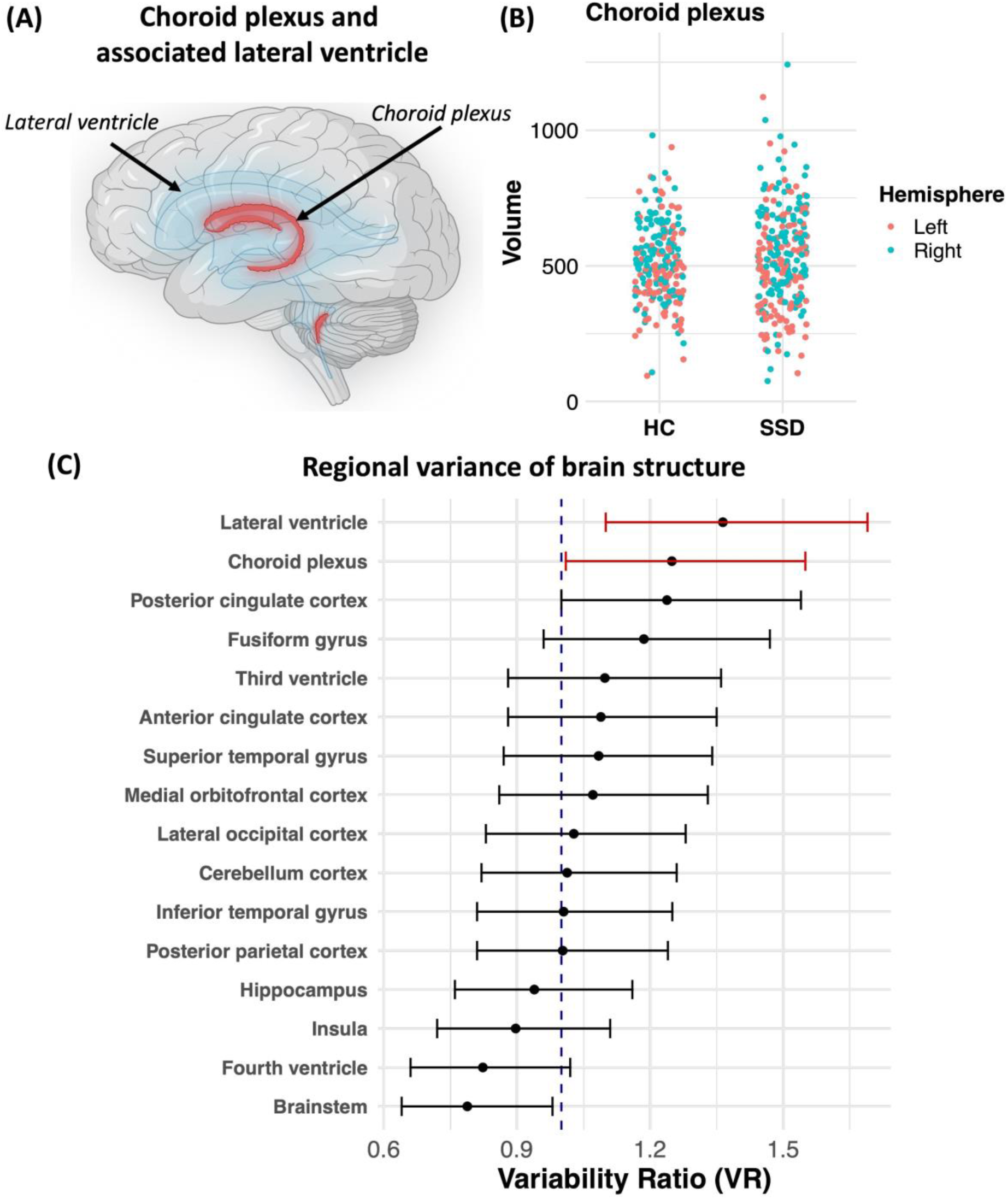
Comparison of choroid plexus volumes and structural variance between individuals with schizophrenia-spectrum disorders and healthy controls. **(A)** Schematic illustration of the choroid plexus (red) and associated lateral ventricle (blue) of the brain. **(B)** Comparison of mean volumes of the left (red) and right (turquoise) choroid plexus between the healthy control (HC) group and the schizophrenia-spectrum disorders (SSD) group, illustrated with a jitter plot. Data points represent individual choroid plexus volumes. Groups were compared using a linear mixed-effect model, controlling for age and sex. N_HC_ = 107, n_HC_ = 214, N_SSD_ = 132, n_SSD_ = 264. **(C)** Forest plot depicting the variability ratio (VR) of choroid plexus, lateral ventricle (red) and other regions of interest, implicated in severe mental illness and schizophrenia (black). N_HC_ = 107, N_SSD_ = 132. Abbreviations: N, number of participants; n, number of choroid plexus volumes; VR, variability ratio. *Created with BioRender.com*.

To validate the automatic segmentation of ChP via FreeSurfer, two independent raters (VY and MK), trained by a neuroimaging expert (DK), manually segmented the ChP in the lateral ventricle on the 3D-T1 images in a subset of 20 randomly selected participants (10 individuals with SSD and 10 HCs), employing ITK-SNAP software, version 4.2.0 (http://www.itksnap.org). The raters were blinded regarding clinical and imaging data and followed a previously published protocol for ChP segmentation^43^. The raters had a high intraclass correlation coefficient for their ratings (0.72) and a moderate Spearman correlation (r = 0.659) with FreeSurfer volumes in line with a previous report^44^ (Figure S1). Details regarding pre- and postprocessing are provided in the Supplementary Methods.

### 2.5 Statistical analyses

The R language (v4.2.1, R Core Team, 2021)^45^ in RStudio environment (RStudio Team, 2020) was used for all statistical analyses and visualizations. The following statistical tests were used to compare differences in demographic and clinical variables between SSD and HC: Fisher’s exact test for categorical variables, Welch’s *t* test for normally distributed and Wilcoxon rank-sum test for non-normally distributed continuous variables. Shapiro – Wilk test was used to assess normality within groups^46^.

Linear mixed-effects models (LMM) based on restricted maximum likelihood estimates were calculated for the association between SSD and choroid plexus volume using the “lme4” package^47^. We included both hemispheres and adjusted for the correlation of the measurements of each participant’s hemisphere by including a random intercept for participant identification number. To explore the relationships with choroid plexus volume as the outcome variable, we incorporated a group variable (SSD vs HC), hemisphere (left vs right), and an interaction term between the group and hemisphere as fixed factors in the model. We compared the volumes between groups and between hemispheres and added an interaction of both to investigate whether a possible group effect is mainly driven by a specific hemisphere. Sex and age were used as covariables due to their impact on choroid plexus volume^19^.

To quantify the variability in the volumes of the regions of interest in SSD patients and HC, we employed the Variability Ratio (VR), a technique adapted from Brugger & Howes^2^.

The logarithm of the variability ratio (lnVR) was determined using the formula:

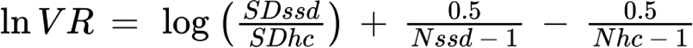

where *SDssd* and *SDhc* are the standard deviations for SSD and HC respectively, and *Nssd* and *Nhc* are the sample sizes for SSD and HC respectively. The effect size for lnVR was transformed back to linear scale using the following formula to aid interpretation of results:

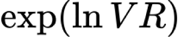

A mean VR > 1, whose 95% confidence interval did not include 1.00 was referred to as increased, as previously suggested^2, 48, 49^.

A K-means clustering analysis within the SSD cohort was conducted based on the choroid plexus and lateral ventricle volume measures to identify subgroups of participants who share similar patterns of ventricular and plexus morphology. Due to larger absolute volumes of the lateral ventricle compared to choroid plexus, we scaled the data, using the *“scale”* function in R, to improve the accuracy of the clustering algorithm^50^. The optimal cluster number (*k*) solution was determined by the elbow method as previously described^51^. Briefly, k-means clustering was performed on the dataset for a range of values for *k*, calculating the within-cluster sum of squares (WSS). The position on the WSS plot, where the slope rapidly decreases (the “elbow”), was selected as the optimal cluster number solution (Fig. S3).

Group effects between clusters were tested using analysis of covariance (ANCOVA), including age and sex as covariates. For differences regarding numbers of neutrophils, monocytes and lymphocytes smoker status was additionally added as a covariate^52^. For differences regarding levels of hsCRP, smoking status and BMI were additionally added as covariates as previously suggested^53^. For differences regarding polygenic risk score for schizophrenia (SZ-PRS), the first five multidimensional scaling (MDS) components were additionally added as covariates to adjust for ancestry. Due to unequal sample sizes between clusters, we employed the Games-Howell nonparametric post-hoc test.

To explore the effects of ChP volume on cortical and subcortical regions, previously linked to peripheral inflammation^16^, we performed univariate linear regressions, controlling for age and sex as covariates.

The tests including peripheral leukocytes, hsCRP, and SZ-PRS were only performed in a subgroup of participants (76, 78, and 83 individuals respectively). Since the complete blood count and the serum analyses were only conducted in inpatients and individuals who agreed to blood sampling respectively, and the missing data was “missing not at random” we decided against imputation.

The threshold for statistical significance was set at *p* value < 0.05. Results from the descriptive statistics and the between-cluster comparisons are shown as mean ± standard deviation (SD). To address the multiple comparison issue, we employed the false discovery rate (FDR) procedure, based on Benjamini-Hochberg method, for correction of the *p* values, which were reported as *q* values.

## 3. Results

### 3.1. Cohort characteristics

The study cohort comprised 132 individuals with SSD (76 in- and 56 outpatients) and 107 HCs, who underwent brain MRI (Table 1). There were no significant differences regarding sex ratio and age between the HC (66% male; 37.05 years) and the SSD group (77% male; 37.33 years) (*p* = 0.111; *p* = 0.721). Individuals with SSD had on average higher BMI values (SSD, mean ± SD = 28.09 ± 5.11; HC, mean ± SD = 23.64 ± 3.70, *p* < 0.001) (Table 1). A strong difference was also evident in the smoking status: 50% of the individuals with SSD were active smokers, compared to 11% of the HCs (*p* < 0.001). The most common diagnosis was schizophrenia (72%), followed by schizoaffective disorder (22%), brief psychotic disorder (5%) and delusional disorder (1%). Mean DUI was 143.68 months (SD = 115.42) and mean duration of untreated psychosis was 22 months (SD = 34.80), although we could only collect data on the DUP in 78 of the participants. The PANSS total scores averaged 54.80 (SD = 15.20), indicating that patients were on average “mildly ill” ^54^. Forty-seven individuals (37%) were either being treated with or reported a history of intake of clozapine in their lifetime, while 80 individuals from the SSD cohort have never received clozapine. Out of the 132 participants with SSD, nine were antipsychotic-naïve.

**Table 1.** Cohort characteristics.

| Demographic characteristics | SSDs |  | HC |  | *p* |
| --- | --- | --- | --- | --- | --- |
|  | *Mean ± SD* | *N* | *Mean ± SD* | *N* |  |
| Age, years | 37.33 ± 11.18 | 132 | 37.05 ± 11.85 | 107 | 0.721<sup>a</sup> |
| BMI | 28.09 ± 5.11 | 128 | 23.64 ± 3.70 | 103 | **<0.001<sup>a</sup>** |
|  | *n (%)* |  | *n (%)* |  | *p* |
| Sex, male:female | 101:31 (77%) |  | 71:36 (66%) |  | 0.111<sup>b</sup> |
| Current smoking, yes:no | 64:63 (50%) | 127 | 11:93 (11%) | 104 | **<0.001<sup>b</sup>** |
| Disease characteristics | *Mean ± SD* | *N* | *Mean ± SD* | *N* | *p* |
| Duration of illness, months | 143.68 ± 115.42 | 128 | – | – | – |
| Duration of untreated psychosis, months | 22.00 ± 34.80 | 78 | – | – | – |
| PANSS positive symptoms | 12.82 ± 4.59 | 128 | 7.34 ± 0.83 | 107 | **<0.001<sup>a</sup>** |
| PANSS negative symptoms | 13.45 ± 5.40 | 128 | 7.41 ± 0.91 | 107 | **<0.001<sup>a</sup>** |
| PANSS general symptoms | 28.52 ± 7.79 | 128 | 16.92 ± 1.45 | 107 | **<0.001<sup>a</sup>** |
| PANSS total score | 54.80 ± 15.20 | 128 | 31.64 ± 2.53 | 107 | **<0.001<sup>a</sup>** |
| CPZeq, mg | 335.87 ± 254.14 | 127 | – | – | – |
| Diagnosis (*DSM-5*) | *N (%)* |  | *N (%)* |  | – |
| Schizophrenia | 95 (72%) |  | – |  | – |
| Schizoaffective disorder | 30 (22%) |  | – |  | – |
| Brief psychotic disorder | 6 (5%) |  | – |  | – |
| Delusional disorder | 1 (1%) |  | – |  | – |

BMI, body mass index; CPZeq, chlorpromazine equivalent doses; HC, healthy control; *N*, number of participants; *p*, *p* value; PANSS, Positive and Negative Syndrome Scale; *SD*, standard deviation; SSD, schizophrenia spectrum disorder; MRI, magnetic resonance imaging.
a - Wilcoxon rank sum test
b - Chi-square test
c - Welch Two Sample t-test

### 3.2. Choroid plexus volume and asymmetry in SSD

First, we aimed at replicating the recently published findings on ChP (Figure 1A) enlargement in SSD^19, 30^. For that reason, we compared the ChP volumes between SSD subjects and HCs (Figure 1B; Table S1a, b). Our analysis showed no significant differences in choroid plexus volumes after controlling for age and sex as covariables (estimate [95% CI] = 19.14 mm^3^ [- 15.37, 53.64]; *p* = 0.362). Furthermore, the right choroid plexus was significantly larger than the left one, independent of phenotype (estimate [95% CI] = 66.41 mm^3^ [43.72, 89.1]; *p* < 0.001). Building on the asymmetry hypothesis in schizophrenia^55^, we compared the choroid plexus asymmetry index (Suppl. Methods) between individuals with SSD and HCs. We did not find significant differences between both groups (Table S1c, Figure S2).

### 3.3. Structural variance of choroid plexus

Driven by the observation of increased scattering of ChP volumes in the SSD group (Figure 1B), we investigated whether volumes of the choroid plexus and other associated regions (Figure 1C) show an increased variability (as indicated by variability ratio or VR) in the SSD group, compared to the HC group. We calculated VR for all regions of interest outlined above and implicated in convergent transdiagnostic or schizophrenia-specific brain networks^41, 42^, using an established method^2^. The variability of the ChP (estimate [95% CI] = 1.249 [1.01, 1.55]) and the associated lateral ventricle (estimate [95% CI] = 1.364 [1.1, 1.69]) was more pronounced in the SSD group compared to the variability in the HC group. Except for the posterior cingulate cortex (estimate [95% CI] = 1.238 [1.0, 1.54]), where we saw a non-significant trend towards increased variance, the variability of the volumes and the cortical thickness of all other regions was not higher in the SSD group compared to the HC group (Table S2a). The VR analyses of the unilaterally computed regions of interest mostly confirmed the results of bilaterally computed regional VRs (Table S2b).

### 3.4. Clusters based on regions with increased variability

Next, we conducted a K-means clustering analysis based on the choroid plexus and lateral ventricle (Lat Vent) volumes (as regions of interest with increased variability) to identify subgroups within the SSD cohort with distinct patterns of the ventricular system. Employing the elbow method, we identified the ideal cluster solution to be 3 (Figure S3). According to the scaled mean volumes of the 2 features (ChP and lateral ventricle) in each cluster, we deduced that cluster 1 (N = 11, scaled centers ChP/LatVent = 1.314, 2.366) was “large ChP-large lateral ventricle cluster”, cluster 2 (N = 65, scaled centers ChP/LatVent = −0.76, −0.549) was “small ChP-small lateral ventricle cluster” and cluster 3 (N = 56, scaled centers ChP/LatVent = 0.624, 0.173) was “intermediate ChP-small lateral ventricle cluster” (Table S3a, Figures 2A, S3A).

**Figure 2.**
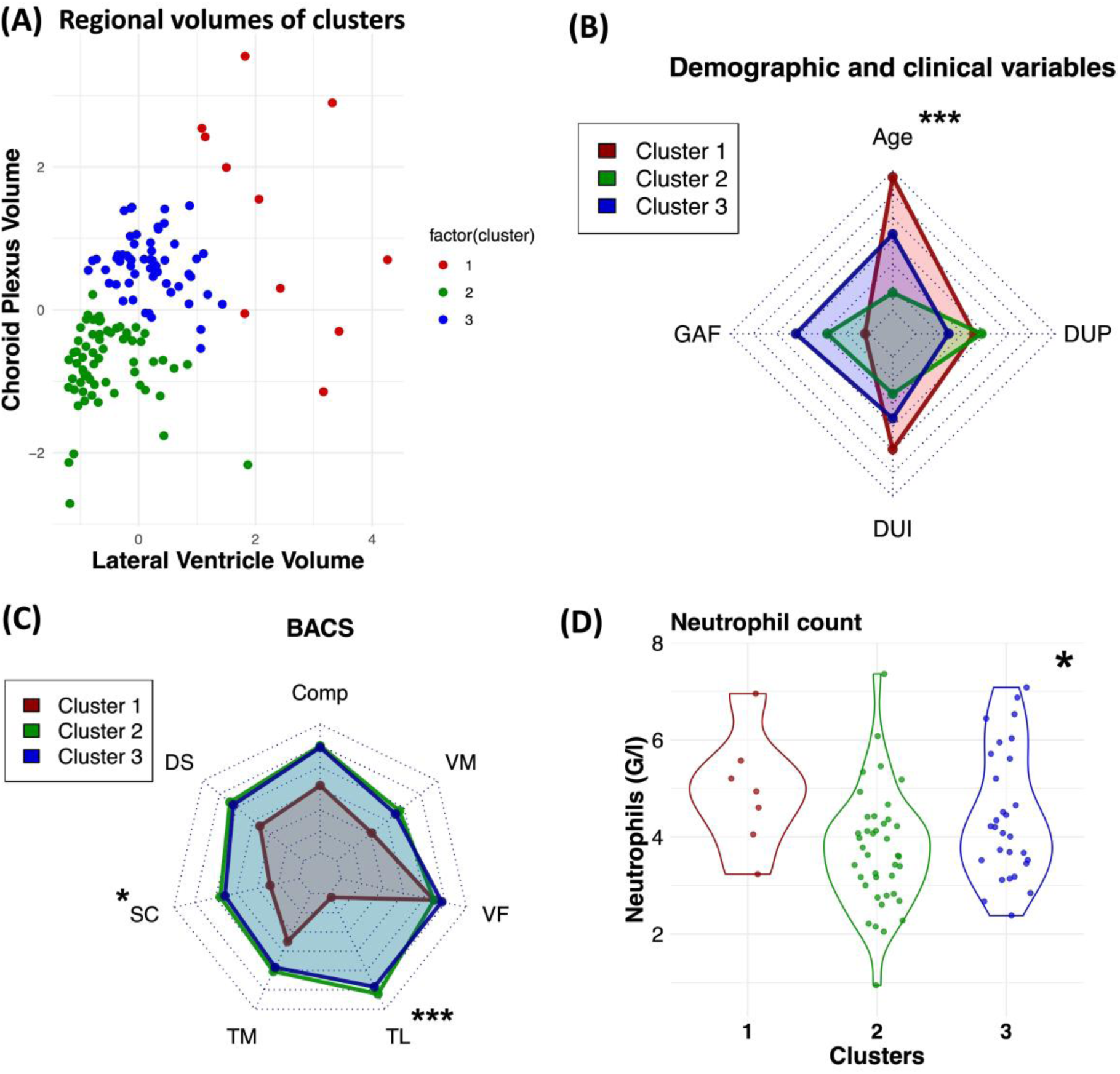
Cluster characteristics. **(A)** Scatter plot depicting the lateral ventricle (x-axis) and choroid plexus (y-axis) volumes of of cluster 1 (red), cluster 2 (green) and cluster 3 (blue) within the SSD cohort. **(B)** Radar chart illustrating cluster mean scaled scores of age, “duration of untreated psychosis” (DUP), “duration of illness” (DUI), global assessment of functioning (GAF). **(C)** Radar chart illustrating cluster mean z-scores of different cognitive subdomains, measured by “The Brief Assessment of Cognition in Schizophrenia” (BACS). **(D)** Comparison of mean peripheral neutrophil counts between cluster 1 (N = 7), cluster 2 (N = 40), and cluster 3 (N = 29). Data points represent individual neutrophil counts. \**q* < 0.05; \*\*\**q* < 0.001. Abbreviations: N, number of participants; DUP, “duration of untreated psychosis”; DUI, “duration of illness”; GAF, “Global Assessment of Functioning”; BACS, “The Brief Assessment of Cognition in Schizophrenia”; Comp, composite score; VM, verbal memory test; VF, verbal fluency test; TL, tower of London test; TM, token motor test; SC, symbol coding test; DS, digit sequence test; G/l, Giga/liter.

### 3.5. Characteristics of clusters within the SSD group

There were no significant across-cluster differences regarding sex (χ^2^ = 2.21, *p* = 0.331), diagnosis (χ^2^ = 2.41, *p* = 0.237), treatment resistance status (χ^2^ = 3.52, *p* = 0.172), and smoking status (χ^2^ = 2.88, *p* = 0.237; Table S3a). We found no significant across-cluster differences regarding DUI, DUP, and BMI (Table S3b, Figures 2B, S4F). There were significant across-cluster differences in terms of age (cluster 1, mean ± SD (years) = 47.7 ± 9.62; cluster 2, mean ± SD = 32.9 ± 9.55, cluster 3, mean ± SD = 40.4 ± 11, *q* < 0.001) and global assessment of functioning (GAF) scores, although the difference in GAF scores did not survive FDR correction (cluster 1, mean ± SD = 46.5 ± 11.9; cluster 2, mean ± SD = 51.6 ± 13.1, cluster 3, mean ± SD = 55.8 ± 8.89, *q* = 0.055; Figure 2B). The Games-Howell post-hoc test showed that the participants from cluster 1 and 3, respectively, were significantly older than those from cluster 2 (p < 0.001; Table S3b).

There were no significant differences regarding psychopathology (operationalized by PANSS) across clusters (Figure S4A, Table S3c). While we didn’t find significant across-cluster differences regarding global cognition (operationalized by BACS composite z-score), there were significant differences for BACS symbol coding (cluster 1, mean ± SD (z-score) = −2.38 ± 1.11; cluster 2, mean ± SD = −1.15 ± 0.92, cluster 3, mean ± SD = −1.26 ± 1.19, *q* = 0.025) and tower of London subtests (cluster 1, mean ± SD (z-score) = −3 ± 3.34; cluster 2, mean ± SD = −0.41 ± 1.05, cluster 3, mean ± SD = −0.6 ± 1.72, *q* = 0.001; Figure 2C, Table S3d). The Games-Howell post-hoc tests showed that cluster 1 had lower symbol coding test scores than cluster 2 (*p* = 0.026) and 3 (*p* = 0.044) respectively but did not find significant between-cluster differences for the tower of London test scores.

Due to the role of the choroid plexus as an immunological niche in the CNS we compared the across-cluster differences regarding peripheral leukocytes and hsCRP levels in two subgroups of participants. We found significant across-cluster differences regarding absolute neutrophil (cluster 1, mean ± SD (G/l) = 4.93 ± 1.18; cluster 2, mean ± SD = 3.72 ± 1.2, cluster 3, mean ± SD = 4.44 ± 1.33, *q* = 0.044) and monocyte (cluster 1, mean ± SD (G/l) = 0.637 ± 0.14; cluster 2, mean ± SD = 0.517 ± 0.145, cluster 3, mean ± SD = 0.496 ± 0.12, *q* = 0.044; Figures 2D, S4B) counts as well as hsCRP levels (cluster 1, mean ± SD (pg/ml) = 3.78×10^8^ ± 3.94×10^8^; cluster 2, mean ± SD = 2.20 ×10^8^ ± 2.35 ×10^8^, cluster 3, mean ± SD = 1.60 ×10^8^ ± 1.49 ×10^8^, *q* = 0.044; Figure S4D). The Games-Howell post-hoc test revealed a trend towards higher neutrophil counts in cluster 3 (*p* = 0.063) and 1 (*p* = 0.083), respectively, compared to cluster 2 that did not reach statistical significance (Table S3e). The Games-Howel post-hoc tests regarding monocytes and hsCRP levels did not show significant between-cluster differences (Table S3e).

Furthermore, we did not find significant across-cluster differences regarding the polygenic risk score for schizophrenia (SZ-PRS) after controlling for age, sex, and the first five MDS components as covariates (Table S3f, Figure S4E).

### 3.5. Relationship between choroid plexus and brain structure

Since the choroid plexus builds the interface between peripheral immunity and CNS we then explored the relationship between choroid plexus volume and the volumes of cortical and subcortical structures, previously associated with peripheral inflammation in SSD^16^ (Table S4a, S4b). Among the subcortical regions of interest we discovered a positive association between choroid plexus volume and volumes of hippocampus (estimate [95% CI] = 1.41 [0.78, 2.03]; *q* = 0.001), thalamus (estimate [95% CI] = 1.93 [0.67, 3.2]; *q* = 0.024), and putamen (estimate [95% CI] = 1.18 [0.37, 2.0]; *q* = 0.024), but not amygdala within the SSD cohort (Table S4a, Figure 3). Furthermore, those associations appeared to be specific for SSD since we did not find significant associations between those structures in the HC group (Table S5, Figure S7). Interestingly, we did not find significant associations between the choroid plexus volume and the volumes of cortical structures, previously associated with peripheral inflammation (Table S4b, Figure S5). Moreover, there was no association between the volumes of hippocampus, thalamus, putamen, and the volume of the other structure with increased variance, namely the lateral ventricle (Table S4c, Figure S6).

**Figure 3.**
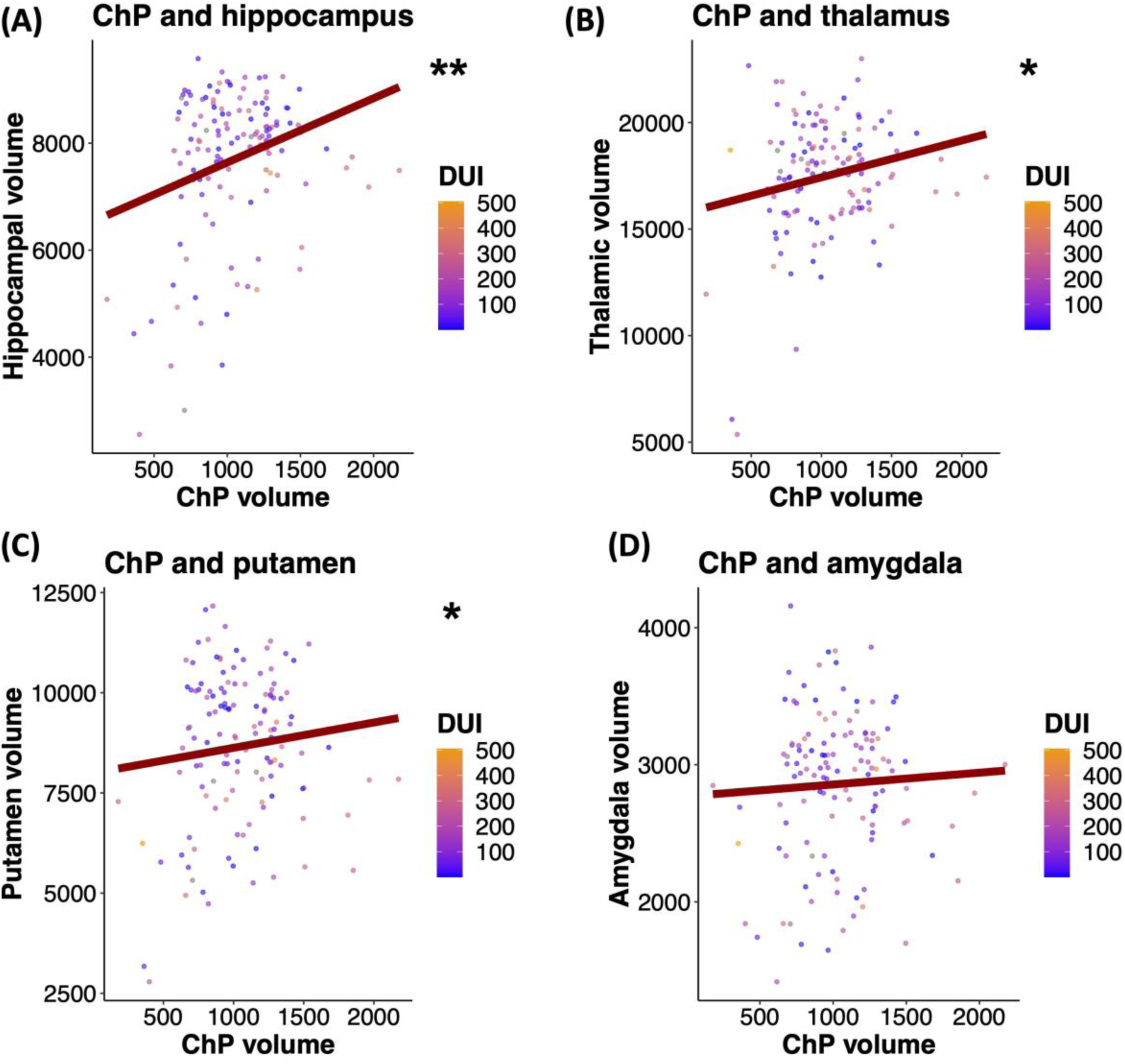
Association between volumes of choroid plexus and subcortical regions, previously linked to peripheral inflammation in SSD. Regression plots illustrating relationships between choroid plexus volume, **(A)** hippocampus, **(B)**, thalamus, **(C)** putamen, and **(D)** amygdala volumes in SSD. The analyses were conducted via univariate linear regressions, controlling for age and sex. Data points represent individual volumes and are colour-coded for “duration of illness” with orange colour indicating higher and blue colour indicating lower duration. N_SSD_ = 132. \**q* < 0.05; \*\**q* < 0.01. Abbreviations: N, number of participants; DUI, “duration of illness (months)”; ChP, choroid plexus.

## 4. Discussion

In this cross-sectional study we could not replicate previous findings^19, 30, 56^ for increased mean volume of the choroid plexus in SSD compared to HCs but observed a clear scattering of the volumes within the SSD group, which prompted us to investigate this variability further. Using a well-established technique for quantification of structural variance^2^, we demonstrated an increased variance of the ChP and the lateral ventricle in SSD as opposed to other regions implicated in SSD and other neuropsychiatric disorders, suggesting a measurable degree of heterogeneity. Employing a K-means clustering analysis, based on the volumes of ChP and lateral ventricle, we identified 3 clusters within the SSD cohort with differences in age, cognitive abilities, and possibly peripheral inflammatory measures. Furthermore, the ChP volume positively correlated with volumes of subcortical, but not cortical regions, previously associated with peripheral inflammation in SSD^16^ and this relationship appeared to be specific for the SSD group.

Some of the possible reasons for the discrepancy between our and previous findings include between-study differences in cohorts (absence of participants with affective psychosis in our study), age differences (our SSD cohort was on average older than the cohort of Zhou et al.)^30,56^ and symptom severity (our cohort had on average less severe psychopathology than the cohorts of Zhou et al. and Lizano et al.)^19, 56^. The high interindividual variability of the ChP volumes within the SSD group together with a lower sample size than Lizano et al.^19^ might have also contributed to the lack of statistical significance in the between-group analysis. Another important factor that might have contributed to the discrepancy between our study and the one of Senay et al. is the difference in segmentation strategy^30^.

While there is some evidence suggesting an increased mean ChP volume, to our knowledge no study so far has investigated the variability of the ChP in any neuropsychiatric disorder. A meta-analysis including measurements from 3901 patients with first-episode schizophrenia and 4040 controls demonstrated increased VR of third ventricle, putamen and thalamus and temporal lobe, decreased VR of the anterior cingulate cortex volume, compared to HCs^2^. In our study we extended this knowledge and found for our SSD sample a significant increase in VR of choroid plexus and lateral ventricle, but not in other regions, implicated in SSD or other neuropsychiatric disorders. The fact that the ChP and the lateral ventricle showed higher variance as opposed to the other regions suggests that alterations of the ventricular system might be seen in a SSD subgroup and could serve as an *intermediate phenotype* for stratification purposes. Taken together, the higher variance of the ChP suggests that it might be enlarged only in a subgroup of people with SSD.

To explore this variance, we conducted a K-means clustering analysis and identified three clusters with distinct patterns of ventricular system within the SSD group. Cluster 1 was characterized by both enlarged choroid plexus and lateral ventricle while cluster 2 had low choroid plexus and lateral ventricle volumes. The participants from cluster 3 showed relatively low lateral ventricle and relatively high choroid plexus volumes. Next, we aimed at investigating how the clusters differ regarding demographic, clinical features as well as peripheral inflammatory markers and genetic liability for schizophrenia. All three clusters differed regarding age with cluster 3 consisting of participants on average older than the ones from cluster 2 and younger than the ones from cluster 1. Those results were in line with previous evidence suggesting that age impacts the volumes of both ChP and lateral ventricle^19, 57^. There were no significant across-cluster differences regarding BMI and smoking status (Table S3a, b), suggesting that they do not significantly contribute to the increased variance of ChP and lateral ventricle. While we did not observe significant differences in psychopathology, measured by PANSS, we saw a trend towards across-cluster differences in level of functioning that did not reach statistical significance. Furthermore, participants from cluster 1 performed significantly worse in the BACS symbol coding test, which measures attention and speed of information processing, than ones from clusters 2 and 3. Interestingly, this data is in line with previous findings, showing an association between choroid plexus volume and worse performance in symbol coding among people with psychosis^19^. Even though we found significant across-cluster differences regarding absolute neutrophil and monocyte counts as well as hsCRP levels in the main analyses, we could not detect significant pairwise differences between the clusters in the post-hoc analyses. These results suggest that while differences in levels of peripheral inflammation across the three cerebroventricular clusters might be evident, our study could have been underpowered to robustly detect specific between-cluster differences. Although our findings are concomitant with a previous study suggesting a link between peripheral inflammation and choroid plexus volume in psychosis^19^, they have to be interpreted carefully due to the limited sample size of our clusters and the lack of statistical robustness. Follow-up well-powered studies are needed to address this limitation.

Furthermore, we did not find across-cluster differences in genetic liability for schizophrenia, captured by SZ-PRS. Our results suggest that genetic risk for schizophrenia, captured by SZ-PRS, does not significantly contribute to the increased variance of cerebroventricular regions in SSD. While the SZ-PRS only captures a small proportion of the complete genetic background of schizophrenia, and our analyses have limited statistical power, it is possible that the increased variability of cerebroventricular measures in SSD are rather associated with environmental factors linked to the disorders. Nevertheless, this hypothesis renders further testing in subsequent studies.

In a next step we focused on the relationship between ChP volume and (sub-)cortical regions, previously associated with peripheral inflammation in SSD^16^. While we did not find any association between ChP volumes and volumes of any cortical region of interest, we observed a positive association between ChP measures and volumes of hippocampus, putamen, and thalamus, but not amygdala. Furthermore, those associations appeared to be specific for SSD, since we did not observe them in our HC cohort. Interestingly, we did not detect any association between the lateral ventricle volumes and any of the regions, previously linked to peripheral inflammation, further strengthening the notion that the associations are specific to ChP and not just a global effect. It is hard to deduce from mere associations of volumetric measures to neurobiological mechanisms behind those volumetric changes. It has been speculated that volumetric increases of regions, associated with peripheral inflammation, could be a result of compromised BBB and/or BCB, resulting in vasodilation, leukodiapedesis and plasma exudation in the brain^16, 58^. Even though we do not know why we only see an association between ChP and the subcortical, but not the cortical regions, previously associated with peripheral inflammation, one can speculate that the physical distance to the ChP might play a role. Since the studied subcortical structures are in direct vicinity to the ChP and the ChP forms the BCB and acts as an invasion route for peripheral leukocytes^23^, it could be possible that it acts as a mediator between peripheral inflammation and brain structure in SSD. Nevertheless, this hypothesis renders further verification in subsequent investigations.

Some of the limitations of our study include its cross-sectional design and the fact that it is based on a monocentric cohort. This is to our knowledge the second largest SSD cohort where ChP alterations have been studied, however, the increased variability ratio of this region requires larger sample sizes to provide conclusive data on its morphological changes in SSD. Even though we showed a moderate correlation between FreeSurfer segmentation and manual segmentation of ChP, some authors have questioned the reliability of FreeSurfer segmentation of ChP and have suggested other more reliable methods^43, 59^. Furthermore, since only nine of the individuals with SSD in our cohort were antipsychotic-naïve, we cannot exclude medication effects on choroid plexus morphology^21^. The individual clusters (especially cluster 1) might have been underpowered to detect differences regarding clinical and immunological features. Another important limitation of our study is the lack of in-depth analyses of peripheral inflammatory markers, which did not allow us to establish a robust link between peripheral inflammation, choroid plexus alterations and volumetric changes of subcortical brain structures in SSD. Even though an association between peripheral inflammation and the (sub-)cortical regions of interest (Table S4, S5) has been assumed, Lizano’s study remains to the best of our knowledge the only one showing such a link and renders replication. Furthermore, the complete blood count is not only a nonspecific measurement of peripheral inflammation, but it was also only assessed in acutely ill inpatients, which limits the generalizability of our findings. Also, some of the variability of the complete blood count could be explained by the fact that even though blood was collected consistently during the morning hours, it was not always done under fasting conditions.

## 5. Conclusions

In conclusion, we could not replicate the findings of previously reported choroid plexus enlargement in SSD but discovered increased structural variance of this region and the lateral ventricle. A clustering analysis based on the choroid plexus and lateral ventricle volumes identified 3 distinct clusters which differed in age, cognitive functioning, and possibly peripheral inflammatory measures. Furthermore, the choroid plexus volume was positively associated with volumes of hippocampus, thalamus, and putamen, which were previously linked to peripheral inflammation in SSD. Overall, our study highlights the importance of replication efforts in external cohorts. Future investigations with larger samples, more thorough immunological characterization and longitudinal design could help elucidate the role of the choroid plexus in the pathophysiology of SSD and assess its utility as an *intermediate phenotype* for patient stratification.

## Data Availability

All data produced in the present work are contained in the manuscript.

## Acknowledgements

The study was endorsed by the Federal Ministry of Education and Research (Bundesministerium für Bildung und Forschung [BMBF]) within the initial phase of the German Center for Mental Health (DZPG) (grant: 01EE2303C to AH and 01EE2303A, 01EE2303F to PF, AS). This research was supported by BMBF with the EraNet project GDNF UpReg (01EW2206) to VY and PF. The procurement of the Prisma 3T MRI scanner was supported by the Deutsche Forschungsgemeinschaft (DFG, INST 86/1739-1 FUGG). No funding was received by commercial or not-for-profit sectors. EB and MSK were supported by doctoral scholarships from the Faculty of Medicine, LMU Munich, Munich, Germany. LK, IM and FJR were supported by the Else Kröner-Fresenius Foundation for the Residency/PhD track of the International Max Planck Research School for Translational Psychiatry (IMPRS-TP), Munich, Germany. FJR, MC, JM were supported by the Förderprogramm für Forschung und Lehre (FöFoLe) of the Faculty of Medicine, LMU Munich, Munich, Germany. ECS was supported by the Munich Clinician Scientist Program (MCSP).

## Contributions

DK, EW, and FJR designed and conceptualized the Clinical Deep Phenotyping Study. VY, DK, and EW designed this study and wrote the protocol. VY, JM, LK, EB, EW, and MSK recruited patients and collected study data. EW, VY and JM trained staff on diagnostic and clinical assessments. MRI measurements were performed by JM and MSK under the supervision of DK and LR. Genotyping and genetic analyses were performed by SP, FJR, ECS and VY. Serological analyses were performed by NG and EM. Statistical analyses were performed by VY. Data visualization was performed by VY. VY wrote the first draft of the manuscript. JM, LR, MSK, EB, MM, SP, LK, MC, ECS, NG, EM, SH, NW, DS, IM, AH, PF, AS, FJR, DK and EW provided critical review. VY prepared the final manuscript version with the help of all authors.

## References

1. Owen MJ, Sawa A, Mortensen PB. Schizophrenia. Lancet Jul 2 2016;388(10039):86–97.

2. Brugger SP, Howes OD. Heterogeneity and Homogeneity of Regional Brain Structure in Schizophrenia: A Meta-analysis. JAMA Psychiatry Nov 1 2017;74(11):1104–1111.

3. Hyman SE. Revolution stalled. Sci Transl Med Oct 10 2012;4(155):155cm111.

4. Owen MJ. New approaches to psychiatric diagnostic classification. Neuron Nov 5 2014;84(3):564–571.

5. Fillman SG, Weickert TW, Lenroot RK, Catts SV, Bruggemann JM, Catts VS, Weickert CS. Elevated peripheral cytokines characterize a subgroup of people with schizophrenia displaying poor verbal fluency and reduced Broca’s area volume. Mol Psychiatry Aug 2016;21(8):1090–1098.

6. Halstead S, Siskind D, Amft M, Wagner E, Yakimov V, Shih-Jung Liu Z, Walder K, Warren N. Alteration patterns of peripheral concentrations of cytokines and associated inflammatory proteins in acute and chronic stages of schizophrenia: a systematic review and network meta-analysis. Lancet Psychiatry Apr 2023;10(4):260–271.

7. Bishop JR, Zhang L, Lizano P. Inflammation Subtypes and Translating Inflammation-Related Genetic Findings in Schizophrenia and Related Psychoses: A Perspective on Pathways for Treatment Stratification and Novel Therapies. Harv Rev Psychiatry Jan-Feb 01 2022;30(1):59–70.

8. Goldsmith DR, Rapaport MH, Miller BJ. A meta-analysis of blood cytokine network alterations in psychiatric patients: comparisons between schizophrenia, bipolar disorder and depression. Mol Psychiatry Dec 2016;21(12):1696–1709.

9. Lizano P, Pong S, Santarriaga S, Bannai D, Karmacharya R. Brain microvascular endothelial cells and blood-brain barrier dysfunction in psychotic disorders. Molecular Psychiatry 2023/09/01 2023;28(9):3698–3708.

10. Campana M, Strauss J, Munz S, et al. Cerebrospinal Fluid Pathologies in Schizophrenia-Spectrum Disorder-A Retrospective Chart Review. Schizophr Bull Jan 21 2022;48(1):47–55.

11. Yakimov V, Moussiopoulou J, Hasan A, Wagner E. The common misconception of blood-brain barrier terminology in psychiatry and neurology. Eur Arch Psychiatry Clin Neurosci Nov 23 2023.

12. Rømer TB, Jeppesen R, Christensen RHB, Benros ME. Biomarkers in the cerebrospinal fluid of patients with psychotic disorders compared to healthy controls: a systematic review and meta-analysis. Mol Psychiatry Jun 2023;28(6):2277–2290.

13. Warren N, O’Gorman C, Horgan I, et al. Inflammatory cerebrospinal fluid markers in schizophrenia spectrum disorders: A systematic review and meta-analysis of 69 studies with 5710 participants. Schizophr Res Apr 2024;266:24–31.

14. Campana M, Yakimov V, Moussiopoulou J, et al. Association of symptom severity and cerebrospinal fluid alterations in recent onset psychosis in schizophrenia-spectrum disorders - An individual patient data meta-analysis. Brain Behav Immun Apr 10 2024;119:353–362.

15. Pollak TA, Drndarski S, Stone JM, David AS, McGuire P, Abbott NJ. The blood-brain barrier in psychosis. Lancet Psychiatry Jan 2018;5(1):79–92.

16. Lizano P, Lutz O, Xu Y, et al. Multivariate relationships between peripheral inflammatory marker subtypes and cognitive and brain structural measures in psychosis. Mol Psychiatry Jul 2021;26(7):3430–3443.

17. Laskaris L, Mancuso S, Shannon Weickert C, et al. Brain morphology is differentially impacted by peripheral cytokines in schizophrenia-spectrum disorder. Brain Behav Immun Jul 2021;95:299–309.

18. Alexandros Lalousis P, Schmaal L, Wood SJ, et al. Inflammatory subgroups of schizophrenia and their association with brain structure: A semi-supervised machine learning examination of heterogeneity. Brain Behav Immun Jul 7 2023;113:166–175.

19. Lizano P, Lutz O, Ling G, et al. Association of Choroid Plexus Enlargement With Cognitive, Inflammatory, and Structural Phenotypes Across the Psychosis Spectrum. Am J Psychiatry Jul 1 2019;176(7):564–572.

20. Kim S, Hwang Y, Lee D, Webster MJ. Transcriptome sequencing of the choroid plexus in schizophrenia. Transl Psychiatry Nov 29 2016;6(11):e964.

21. Williams MR, Macdonald CM, Turkheimer FE. Histological examination of choroid plexus epithelia changes in schizophrenia. Brain Behav Immun Jul 2023;111:292–297.

22. Falcao AM, Marques F, Novais A, Sousa N, Palha JA, Sousa JC. The path from the choroid plexus to the subventricular zone: go with the flow! Front Cell Neurosci 2012;6:34.

23. Castellani G, Croese T, Peralta Ramos JM, Schwartz M. Transforming the understanding of brain immunity. Science Apr 7 2023;380(6640):eabo7649.

24. Saunders NR, Dziegielewska KM, Fame RM, Lehtinen MK, Liddelow SA. The choroid plexus: a missing link in our understanding of brain development and function. Physiol Rev Jan 1 2023;103(1):919–956.

25. Yakimov V, Schweiger F, Zhan J, Behrangi N, Horn A, Schmitz C, Hochstrasser T, Kipp M. Continuous cuprizone intoxication allows active experimental autoimmune encephalomyelitis induction in C57BL/6 mice. Histochem Cell Biol Aug 2019;152(2):119–131.

26. Borlongan CV, Thanos CG, Skinner SJ, Geaney M, Emerich DF. Transplants of encapsulated rat choroid plexus cells exert neuroprotection in a rodent model of Huntington’s disease. Cell Transplant 2008;16(10):987–992.

27. Klistorner S, Barnett MH, Parratt J, Yiannikas C, Graham SL, Klistorner A. Choroid plexus volume in multiple sclerosis predicts expansion of chronic lesions and brain atrophy. Ann Clin Transl Neurol Oct 2022;9(10):1528–1537.

28. Bergsland N, Dwyer MG, Jakimovski D, Tavazzi E, Benedict RHB, Weinstock-Guttman B, Zivadinov R. Association of Choroid Plexus Inflammation on MRI With Clinical Disability Progression Over 5 Years in Patients With Multiple Sclerosis. Neurology Feb 28 2023;100(9):e911–e920.

29. Assogna M, Premi E, Gazzina S, et al. Association of Choroid Plexus Volume With Serum Biomarkers, Clinical Features, and Disease Severity in Patients With Frontotemporal Lobar Degeneration Spectrum. Neurology Jul 27 2023.

30. Senay O, Seethaler M, Makris N, et al. A preliminary choroid plexus volumetric study in individuals with psychosis. Hum Brain Mapp Apr 15 2023;44(6):2465–2478.

31. Krcmar L, Jager I, Boudriot E, et al. The multimodal Munich Clinical Deep Phenotyping study to bridge the translational gap in severe mental illness treatment research. Front Psychiatry 2023;14:1179811.

32. Kalman JL, Burkhardt G, Adorjan K, et al. Biobanking in everyday clinical practice in psychiatry—The Munich Mental Health Biobank. Front Psychiatry 2022-July-22 2022;13.

33. Sheehan DV, Lecrubier Y, Sheehan KH, et al. The Mini-International Neuropsychiatric Interview (M.I.N.I.): the development and validation of a structured diagnostic psychiatric interview for DSM-IV and ICD-10. J Clin Psychiatry 1998;59 Suppl 20:22–33;quiz 34-57.

34. Boudriot E, Schworm B, Slapakova L, et al. Optical coherence tomography reveals retinal thinning in schizophrenia spectrum disorders. Eur Arch Psychiatry Clin Neurosci Apr 2023;273(3):575–588.

35. Kay SR, Fiszbein A, Opler LA. The Positive and Negative Syndrome Scale (PANSS) for Schizophrenia. Schizophr Bull 1987;13(2):261–276.

36. Leucht S, Samara M, Heres S, Davis JM. Dose Equivalents for Antipsychotic Drugs: The DDD Method. Schizophr Bull Jul 2016;42 Suppl 1:S90–94.

37. Ruderfer DM, Charney AW, Readhead B, et al. Polygenic overlap between schizophrenia risk and antipsychotic response: a genomic medicine approach. Lancet Psychiatry Apr 2016;3(4):350–357.

38. Keefe RSE, Goldberg TE, Harvey PD, Gold JM, Poe MP, Coughenour L. The Brief Assessment of Cognition in Schizophrenia: reliability, sensitivity, and comparison with a standard neurocognitive battery. Schizophrenia Research 2004/06/01/ 2004;68(2):283–297.

39. Fischl B, Salat DH, Busa E, et al. Whole brain segmentation: automated labeling of neuroanatomical structures in the human brain. Neuron Jan 31 2002;33(3):341–355.

40. Jernigan TL, Zatz LM, Moses JA, Jr., Berger PA. Computed tomography in schizophrenics and normal volunteers. I. Fluid volume. Arch Gen Psychiatry Jul 1982;39(7):765–770.

41. Taylor JJ, Lin C, Talmasov D, et al. A transdiagnostic network for psychiatric illness derived from atrophy and lesions. Nat Hum Behav Mar 2023;7(3):420–429.

42. Opel N, Goltermann J, Hermesdorf M, Berger K, Baune BT, Dannlowski U. Cross-Disorder Analysis of Brain Structural Abnormalities in Six Major Psychiatric Disorders: A Secondary Analysis of Mega- and Meta-analytical Findings From the ENIGMA Consortium. Biol Psychiatry Nov 1 2020;88(9):678–686.

43. Bannai D, Cao Y, Keshavan M, Reuter M, Lizano P. Manual Segmentation of the Human Choroid Plexus Using Brain MRI. J Vis Exp Dec 15 2023(202).

44. Li Y, Zhou Y, Zhong W, et al. Choroid Plexus Enlargement Exacerbates White Matter Hyperintensity Growth through Glymphatic Impairment. Ann Neurol Jul 2023;94(1):182–195.

45. Team RC. R: A language and environment for statistical computing. *R Foundation for Statistical Computing, Vienna*, Austria 2022.

46. du Prel JB, Rohrig B, Hommel G, Blettner M. Choosing statistical tests: part 12 of a series on evaluation of scientific publications. Dtsch Arztebl Int May 2010;107(19):343–348.

47. Bates D MM, Bolker B, Walker S. Fitting Linear Mixed-Effects Models Using lme4. Journal of Statistical Software 2015;67(1):1–48.

48. Cumming G. The new statistics: why and how. Psychol Sci Jan 2014;25(1):7–29.

49. Coulson M, Healey M, Fidler F, Cumming G. Confidence intervals permit, but do not guarantee, better inference than statistical significance testing. Front Psychol 2010;1:26.

50. Lee J, Rizzo S, Altshuler L, Glahn DC, Miklowitz DJ, Sugar CA, Wynn JK, Green MF. Deconstructing Bipolar Disorder and Schizophrenia: A cross-diagnostic cluster analysis of cognitive phenotypes. J Affect Disord Feb 2017;209:71–79.

51. Liu F, Yang D, Liu Y, et al. Use of latent profile analysis and k-means clustering to identify student anxiety profiles. BMC Psychiatry Jan 5 2022;22(1):12.

52. Asthana A, Johnson HM, Piper ME, Fiore MC, Baker TB, Stein JH. Effects of smoking intensity and cessation on inflammatory markers in a large cohort of active smokers. Am Heart J Sep 2010;160(3):458–463.

53. Fernandes BS, Steiner J, Bernstein HG, et al. C-reactive protein is increased in schizophrenia but is not altered by antipsychotics: meta-analysis and implications. Mol Psychiatry Apr 2016;21(4):554–564.

54. Leucht S, Kane JM, Etschel E, Kissling W, Hamann J, Engel RR. Linking the PANSS, BPRS, and CGI: clinical implications. Neuropsychopharmacology Oct 2006;31(10):2318–2325.

55. Schijven D, Postema MC, Fukunaga M, et al. Large-scale analysis of structural brain asymmetries in schizophrenia via the ENIGMA consortium. Proc Natl Acad Sci U S A Apr 4 2023;120(14):e2213880120.

56. Zhou YF, Huang JC, Zhang P, et al. Choroid Plexus Enlargement and Allostatic Load in Schizophrenia. Schizophr Bull Apr 10 2020;46(3):722–731.

57. Scahill RI, Frost C, Jenkins R, Whitwell JL, Rossor MN, Fox NC. A longitudinal study of brain volume changes in normal aging using serial registered magnetic resonance imaging. Arch Neurol Jul 2003;60(7):989–994.

58. Kamintsky L, Cairns KA, Veksler R, Bowen C, Beyea SD, Friedman A, Calkin C. Blood-brain barrier imaging as a potential biomarker for bipolar disorder progression. Neuroimage Clin 2020;26:102049.

59. Tadayon E, Moret B, Sprugnoli G, Monti L, Pascual-Leone A, Santarnecchi E. Improving Choroid Plexus Segmentation in the Healthy and Diseased Brain: Relevance for Tau-PET Imaging in Dementia. J Alzheimers Dis 2020;74(4):1057–1068.

