## Supplementary material for "Investigation of choroid plexus variability in schizophrenia-spectrum disorders – insights from a multimodal study": 2024-06-06-Supplemental_Yakimov et al_Investigation of choroid plexus variability in schizophrenia-spectrum disorders ΓÇô insights from a multimodal study_jpeg.docx

Vladislav Yakimov^1,2,8#^ (ORCID-iD: 0000-0001-9559-7492), Joanna Moussiopoulou^1,3,8^ (ORCID-iD: 0000-0002-0157-6197), Lukas Roell^1,3,8^ (ORCID-iD: 0000-0002-0284-2290), Marcel S. Kallweit^1^, Emanuel Boudriot^1,9^ (ORCID-iD: 0000-0001-6083-6318), Matin Mortazavi^1,4^, Sergi Papiol^7,9^, Lenka Krčmář^1,2,3^, Mattia Campana^1^, Eva C. Schulte^7,12,13^ (ORCID:0000-0003-3105-5672), Nicolas Glaichenhaus^14^ (ORCID: 0000-0002-8997-0470), Emanuela Martinuzzi^14^ (0000-0001-5832-951X), Sean Halstead^5^ (ORCID-iD: 0009-0000-4890-3506), Nicola Warren^5,6^ (ORCID-iD: [0000-0002-0805-1182](https://orcid.org/0000-0002-0805-1182)), Dan Siskind^5,6^ (ORCID-iD: [0000-0002-2072-9216](https://orcid.org/0000-0002-2072-9216)), Isabel Maurus^1,2^, Alkomiet Hasan^4,8^, Peter Falkai^1,8,9^ (ORCID: 0000-0003-2873-8667), Andrea Schmitt^1,8,10^ (ORCID: 0000-0002-5426-4023), Florian Raabe^1,2,9^, CDP Working Group^1,4,9^ , Daniel Keeser^1,3,8*^(ORCID: 0000-0002-0244-1024), Elias Wagner^1,4,8,11*^

^1^Department of Psychiatry and Psychotherapy, LMU University Hospital, LMU Munich, Munich, Germany

^2^International Max Planck Research School for Translational Psychiatry (IMPRS-TP), 80804 Munich, Germany

^3^NeuroImaging Core Unit Munich (NICUM), LMU University Hospital, LMU Munich, 80336 Munich, Germany

^4^Department of Psychiatry, Psychotherapy and Psychosomatics, Faculty of Medicine, University of Augsburg, 86156 Augsburg, Germany

^5^Medical School, The University of Queensland, Brisbane, QLD, Australia

^6^Metro South Addiction and Mental Health, Brisbane, QLD, Australia

^7^Institute of Psychiatric Phenomics and Genomics, LMU University Hospital, LMU Munich, Munich, Germany.

^8^DZPG (German Center for Mental Health), partner site Munich/Augsburg

^9^Max Planck Institute of Psychiatry, 80804 Munich, Germany

^10^Laboratory of Neuroscience (LIM27), Institute of Psychiatry, University of Sao Paulo, 05403-903 São Paulo, Brazil, SP, Brazil

^11^Evidence-based Psychiatry and Psychotherapy, Faculty of Medicine, University of Augsburg, Stenglinstrasse 2, 86156 Augsburg, Germany

^12^Institute of Human Genetics, University Hospital, Faculty of Medicine, University of Bonn, 53127 Bonn, Germany

^13^Department of Psychiatry and Psychotherapy, University Hospital, Faculty of Medicine, University of Bonn, 53127 Bonn, Germany

^14^Institut de Pharmacologie Moléculaire et Cellulaire, Université Côte d'Azur, Centre National de La Recherche Scientifique, Valbonne, France.

CDP Working Group: Valéria de Almeida, Stephanie Behrens, Emanuel Boudriot, Mattia Campana, Fanny Dengl, Peter Falkai, Laura E. Fischer, Nadja Gabellini, Vanessa Gabriel, Thomas Geyer, Katharina Hanken, Alkomiet Hasan, Genc Hasanaj, Alexandra Hirsch, Georgios Ioannou, Iris Jäger, Sylvia de Jonge, Marcel S. Kallweit, Temmuz Karali, Susanne Karch, Berkhan Karslı, Daniel Keeser, Christoph Kern, Nicole Klimas, Maxim Korman, Lenka Krčmář, Isabel Lutz, Julian Mechler, Verena Meisinger, Matin Mortazavi, Joanna Moussiopoulou, Karin Neumeier, Frank Padberg, Boris Papazov, Sergi Papiol, Pauline Pingen, Oliver Pogarell, Siegfried Priglinger, Florian J. Raabe, Lukas Roell, Moritz J. Rossner, Andrea Schmitt, Susanne Schmölz, Enrico Schulz, Benedikt Schworm, Elias Wagner, Sven Wichert, Vladislav Yakimov, Peter Zill

* These authors contributed equally

**#Corresponding author:**

Dr. med. Vladislav Yakimov

Address: Department of Psychiatry and Psychotherapy, University Hospital, LMU Munich, Nussbaumstrasse 7, 80336 Munich, Germany

**Running title:** Choroid plexus alterations in schizophrenia

### Supplemental Methods

#### Quality control of structural MRI data

Quality control for structural MRI data was performed as previously described by our working group^1^. Specifically, a visual inspection and utilization of the quality control software MRIQC^2^ were conducted. Manual assessment included rating overall image quality and evaluating specific sequence-related metrics such as signal-to-noise ratio (SNR), contrast-to-noise ratio (CNR), coefficient of joint variation (CJV), entropy-focused criterion (EFC), foreground-background energy ratio (FBER), median intensity non-uniformity (INU), Full Width at Half Maximum (FWHM), framewise displacement (FD), temporal SNR, and spatial root mean square after temporal differencing (DVARS). Images were flagged for exclusion or for further inspection if they received inadequate manual quality ratings and/or exhibited at least one abnormal quality metric. After image processing, we visually inspected the FreeSurfer segmentations and excluded cases with segmentation errors. A total of two participants were excluded based on criteria related to insufficient image quality.

**Structural MRI data processing**

The structural isotropic 3D T1-weighted MR images underwent processing using FreeSurfer v7.2, employing *recon-all,* as previously described^1^. This processing included motion correction and averaging^3^, non-brain tissue removal^4^, automated Talairach transformation, segmentation of subcortical white and grey matter volumes^5, 6^, intensity normalization^7^, tessellation of the grey matter-white matter boundary, automated topology correction^8, 9^, and surface deformation^10-12^. Detailed information on the FreeSurfer pipeline is available at <http://surfer.nmr.mgh.harvard.edu/>. Regional measures of brain volumes and cortical thickness were extracted from the FreeSurfer output. To account for variations in intracranial volume across all subjects, volumes were corrected using the proportions method^13^.

**Genotyping, quality control and imputation**

Genomic DNA from a subgroup of participants was isolated from venous whole blood and underwent genotyping using Illumina’s Global Screening Array (GSA) v3.0 at Life & Brain GmbH, Bonn, Germany. Illumina GenomeStudio v2.0.4 (Illumina, San Dieago, USA) was used to process GSA genotyping data and obtain genotype calls for the participants of the study. Quality control included single nucleotide polymorphism (SNP) call rate > 98%, individual call rate >98%, minor allele frequency (MAF) > 0.05%, Hardy-Weinberg equilibrium (p > 0.001). For the individual-level quality control data was removed to exclude duplicates, sex-mismatches, heterozygosity rate exceeding 3.90 standard deviations, or a non-European ancestry according to a multidimensional scaling (MDS) analysis. Those steps were conducted using PLINK v1.9/v2 ([www.cog-genomics.org/plink/1.9/)66](http://www.cog-genomics.org/plink/1.9/)66), as previously described^14^. Ancestry variations among participants were assessed using PLINK 1.9. Multidimensional scaling was performed on a pruned subset of approximately ~70,000 to ~90,000 autosomal SNPs, with exclusion of regions with high linkage disequilibrium. Only participants, who clustered with 1000 Genomes Project reference EUR population, were included in the study.

Imputation was performed using the Michigan Imputation Server^15^ with the Haplotype Reference Consortium (HRC) dataset as a reference panel. We performed a post-imputation QC to exclude SNPs that had an imputation quality score of R2 < 0.3, or a MAF < 1%. 7,948,189 SNPs survived post-imputation QC and were available for polygenic risk score (PRS) computation.

**Computation of polygenic risk score**

Next, polygenic risk score for schizophrenia (SZ-PRS) was computed for 83 individuals with SSD using summary statistics from the latest published schizophrenia genome-wide association study (GWAS) (PGC-SCZ3)^16^ (data available at <https://www.med.unc.edu/pgc/results-and-downloads>). For this purpose, effect sizes of posterior single nucleotide polymorphism were inferred under continuous shrinkage priors using PRS-CS^17^, as previously described^18^. A fully Bayesian approach with genotype dosage data was used to estimate the global shrinkage parameter (ϕ)^17^.

**Asymmetry index calculation and statistical analysis**

For the bilaterally paired choroid plexus, we computed the Asymmetry Index (AI)^19^, using the widely-accepted^19, 20^ formula:

$$AI =\frac{L-R}{(L+R)/2}$$

where *L* and *R* represent the left and right hemispheric measurement respectively. It standardizes the AI by adjusting it to the overall size of the bilateral measurement of the choroid plexus. A negative AI value reflects a larger right hemispheric measurement (R > L) and a positive AI value a larger left hemispheric measurement (L > R). Group differences (SSD vs HC) were examined for the choroid plexus, employing a univariate linear regression. The case-control status was included as a predictor variable, the AI as an outcome variable, and age as well as sex as covariates in the model.

### Supplemental Figure Legends

#### Figure S1. Parcellation of the choroid plexus.

T1-weighted image of the brain, depicting the choroid plexus in the lateral ventricles including manual (red and blue) and FreeSurfer (turquoise) segmentations.

**Figure S2.** **Choroid plexus asymmetry in SSD and HC.**

Comparison of mean asymmetry indices of the choroid plexus between the healthy control (HC) and the schizophrenia-spectrum disorders (SSD) group, illustrated with a violin/jitter plot. Data points represent individual choroid plexus asymmetry indices using a univariate linear regression, controlling for age and sex. N_SSD_ = 132, N_HC_ = 107. Abbreviations: N, number of participants.

**Figure S3.** **K-means clustering within the SSD cohort.**

**(A)** Scatter plot of a 3-cluster solution from a K-means cluster analysis. The x-axis and y-axis indicate the first 2 principal components of the clustering analysis. **(B)** Elbow method on within-cluster sum of squares (WSS) plot to determine the optimal clustering solution. The slope of WSS rapidly decreases at a 4-cluster solution, indicating a 3-cluster solution as the optimal for our data. N = 132. Abbreviations: N, number of participants; PC, principal component.

**Figure S4**. **Cluster characteristics.**

**(A)** Radar chart illustrating cluster mean scaled PANSS scores of its subdomains (Pos, Neg, Gen) as well as the sum of those (Total). Comparison of mean peripheral **(B)** monocyte, **(C)** lymphocyte counts, (D) high-sensitive C-reactive protein levels, (E) schizophrenia polygenic risk scores, and body mass index (BMI) between cluster 1 (N_cells_ = 7; N_hsCRP_ = 7; N_PRS-SZ_ = 5), cluster 2 (N_cells_ = 40; N_hsCRP_ = 36; N_PRS-SZ_ = 36), and cluster 3 (N_cells_ = 29; N_hsCRP_ = 35; N_PRS-SZ_ = 42). Data points represent individual (A) – (C) cell counts, (D) hsCRP levels, (E) SZ polygenic risk scores, (F) and BMI values. Abbreviations: N, number of participants; PANSS, “Positive and Negative Syndrom Scale”; Pos, positive symptoms; Neg, negative symptoms; Gen, general symptoms; G/l, Giga/liter; hsCRP, high-sensitive C-reactive protein; PRS-SZ, polygenic risk score for schizophrenia; BMI, body mass index.

**Figure S5.** **Relationship between volumes of choroid plexus and cortical regions, previously linked to peripheral inflammation in SSD.**

Regression plots illustrating the relationships between choroid plexus volume, **(A)** superior temporal gyrus, **(B)**, superior frontal gyrus, **(C)** precentral gyrus, **(D)** fusiform gyrus, and **(E)** caudal middle frontal gyrus volumes in SSD. The analyses were conducted via univariate linear regressions, controlling for age and sex. Data points represent individual volumes and are colour-coded for “duration of illness” with orange colour indicating higher and blue colour indicating lower duration. N_SSD_ = 132. Abbreviations: N, number of participants; DUI, “duration of illness (months)”; ChP, choroid plexus; STG, superior temporal gyrus; SFG, superior frontal gyrus; CMF, caudal middle frontal gyrus.

**Figure S6.** **Relationship between volumes of lateral ventricle and subcortical regions, previously linked to peripheral inflammation in SSD.**

Regression plots illustrating relationships between lateral ventricle volume, **(A)** hippocampus, **(B)**, thalamus, and **(C)** putamen volumes in SSD. The analyses were conducted via univariate linear regressions, controlling for age and sex. Data points represent individual volumes and are colour-coded for “duration of illness” with orange colour indicating higher and blue colour indicating lower duration. N_SSD_ = 132. Abbreviations: N, number of participants; DUI, “duration of illness (months)”; Lat Vent, lateral ventricle.

**Figure S7.** **Association between volumes of choroid plexus and subcortical regions, previously linked to peripheral inflammation in HC.**

Regression plots illustrating relationships between choroid plexus volume, **(A)** hippocampus, **(B)**, thalamus, and **(C)** putamen volumes in HC. The analyses were conducted via univariate linear regressions, controlling for age and sex. Data points represent individual volumes and are colour-coded for age with orange colour indicating higher and blue colour indicating lower age. N_SSD_ = 107. Abbreviations: N, number of participants; ChP, choroid plexus.

**Supplemental Figures**

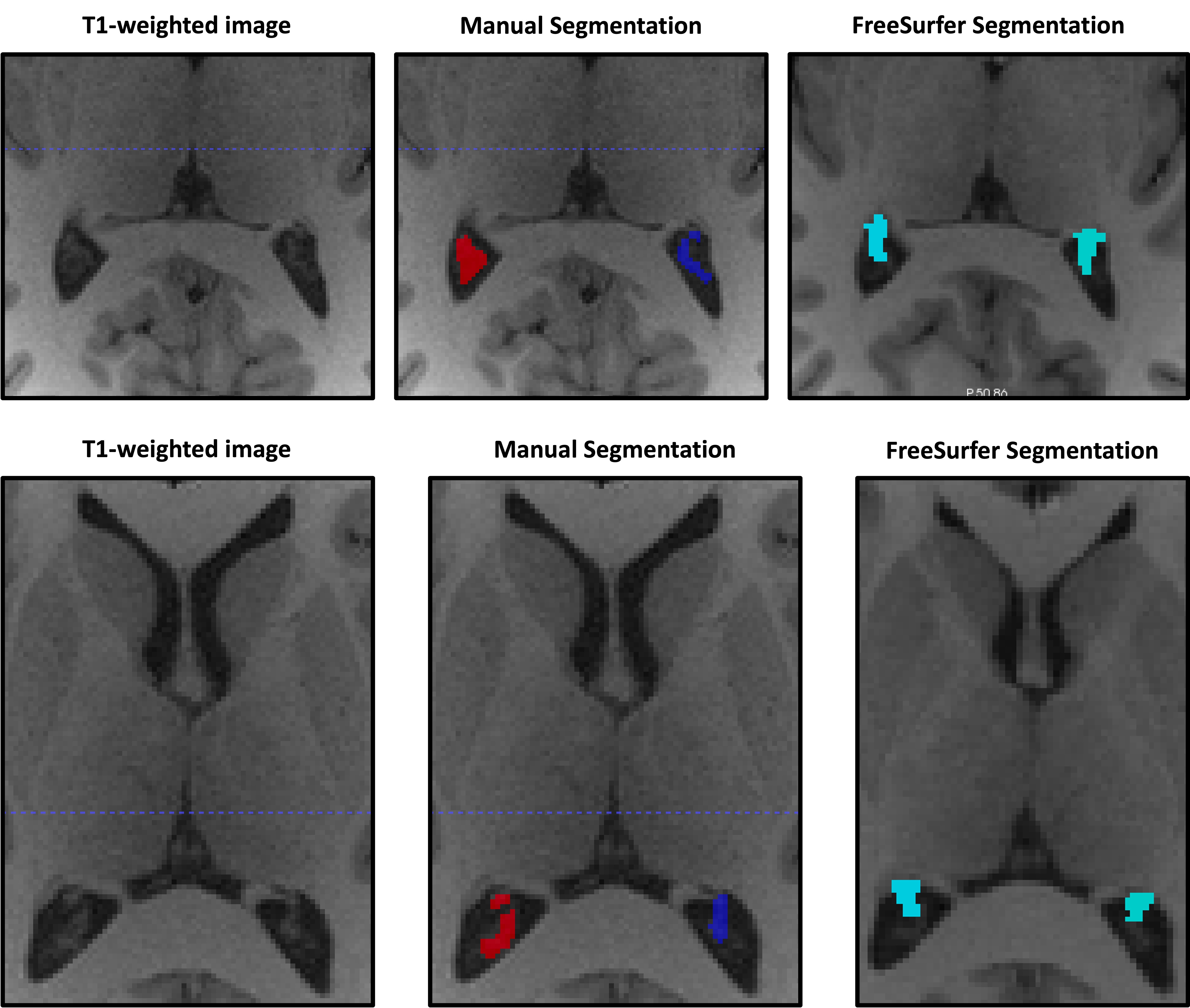

**Figure S1**

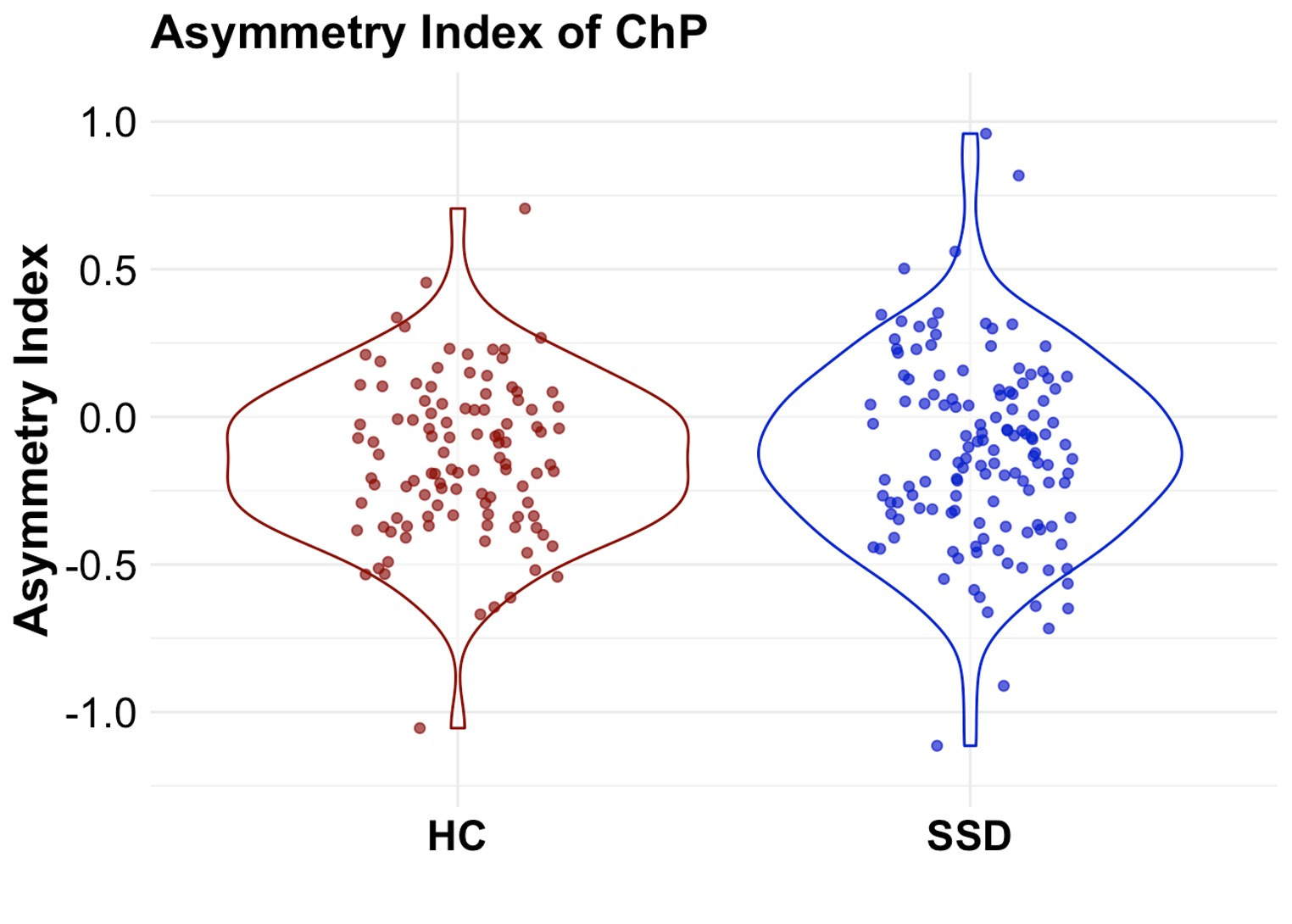

**Figure S2**

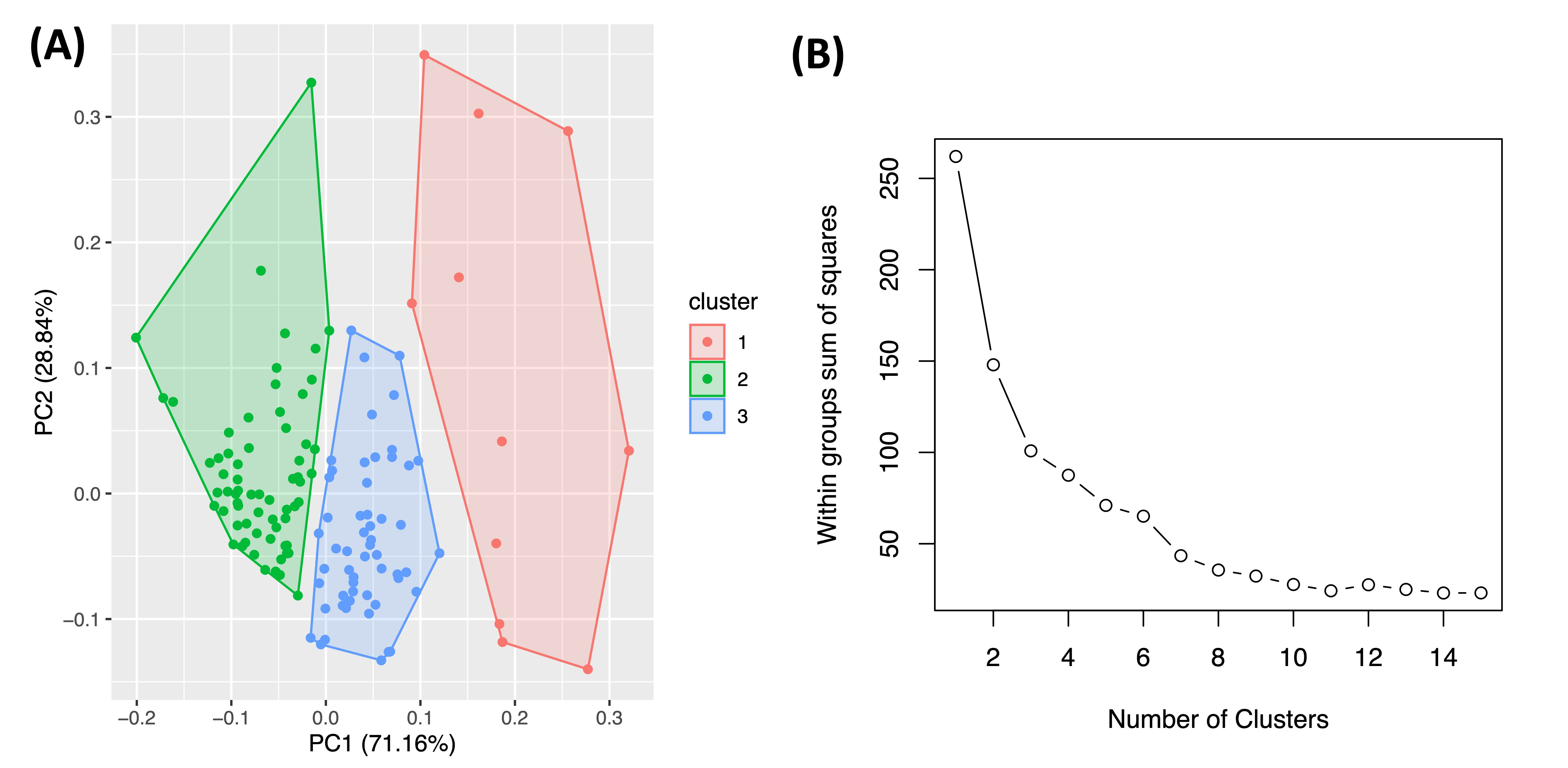

**Figure S3**

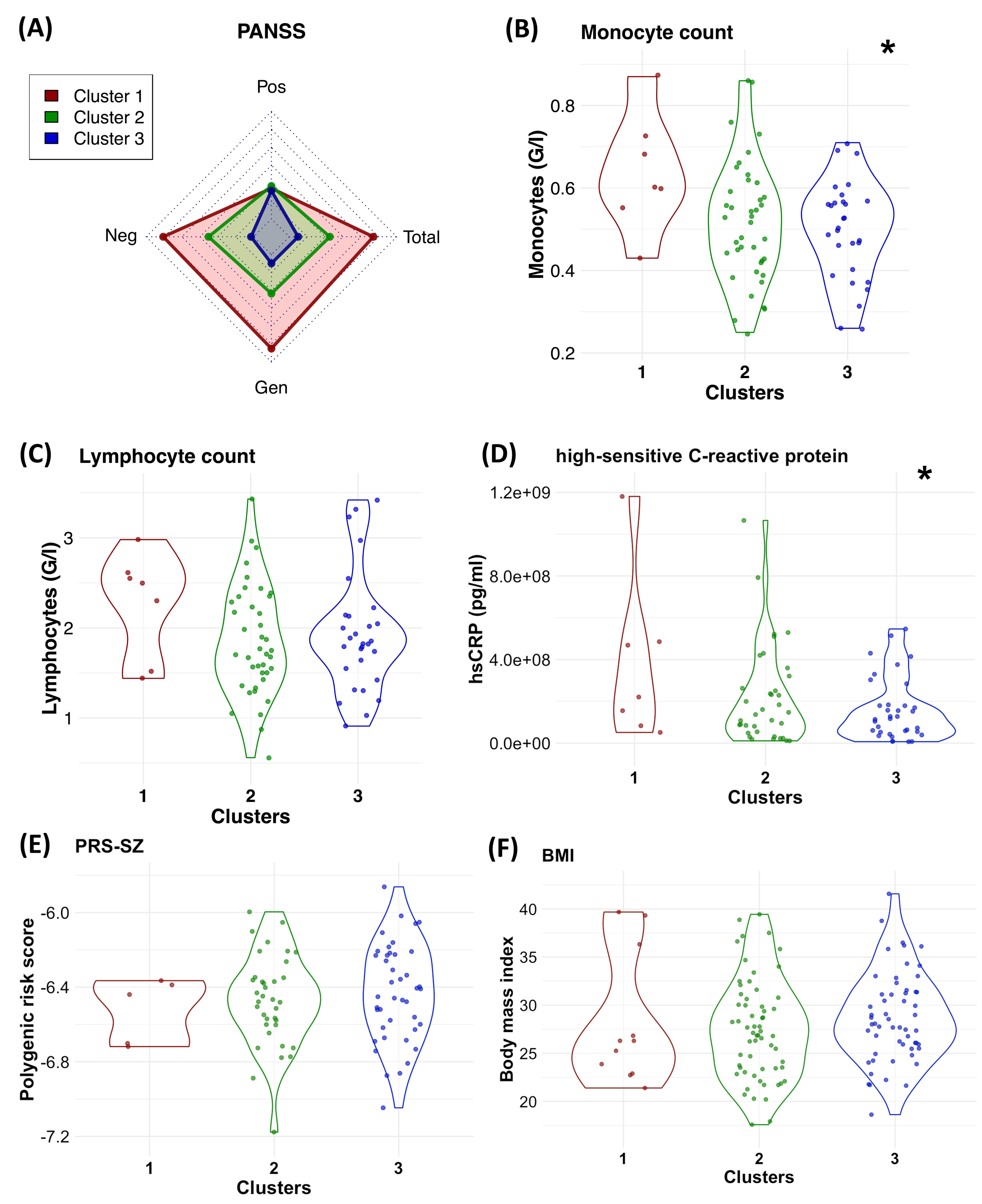

**Figure S4**

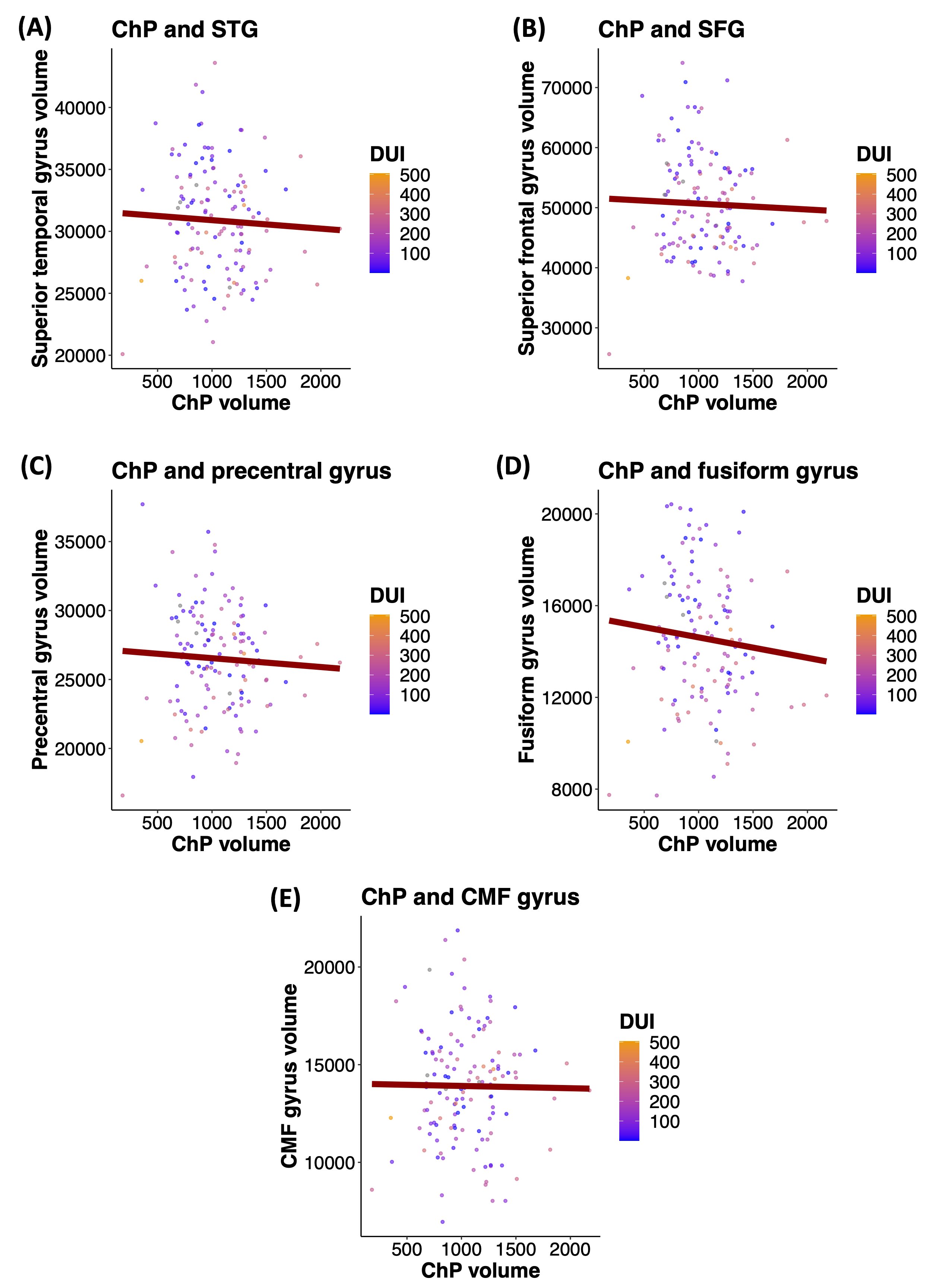

**Figure S5**

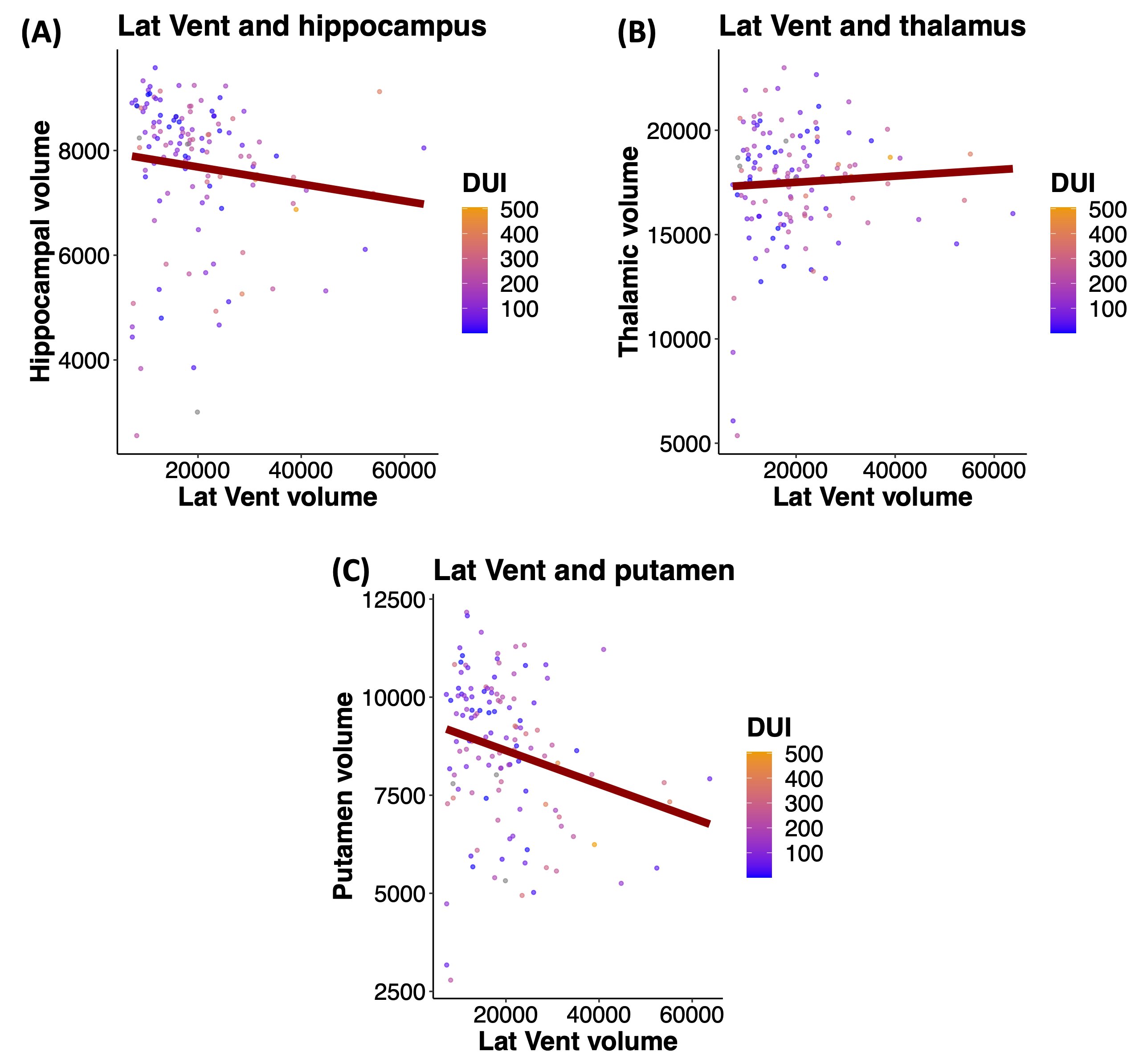

**Figure S6**

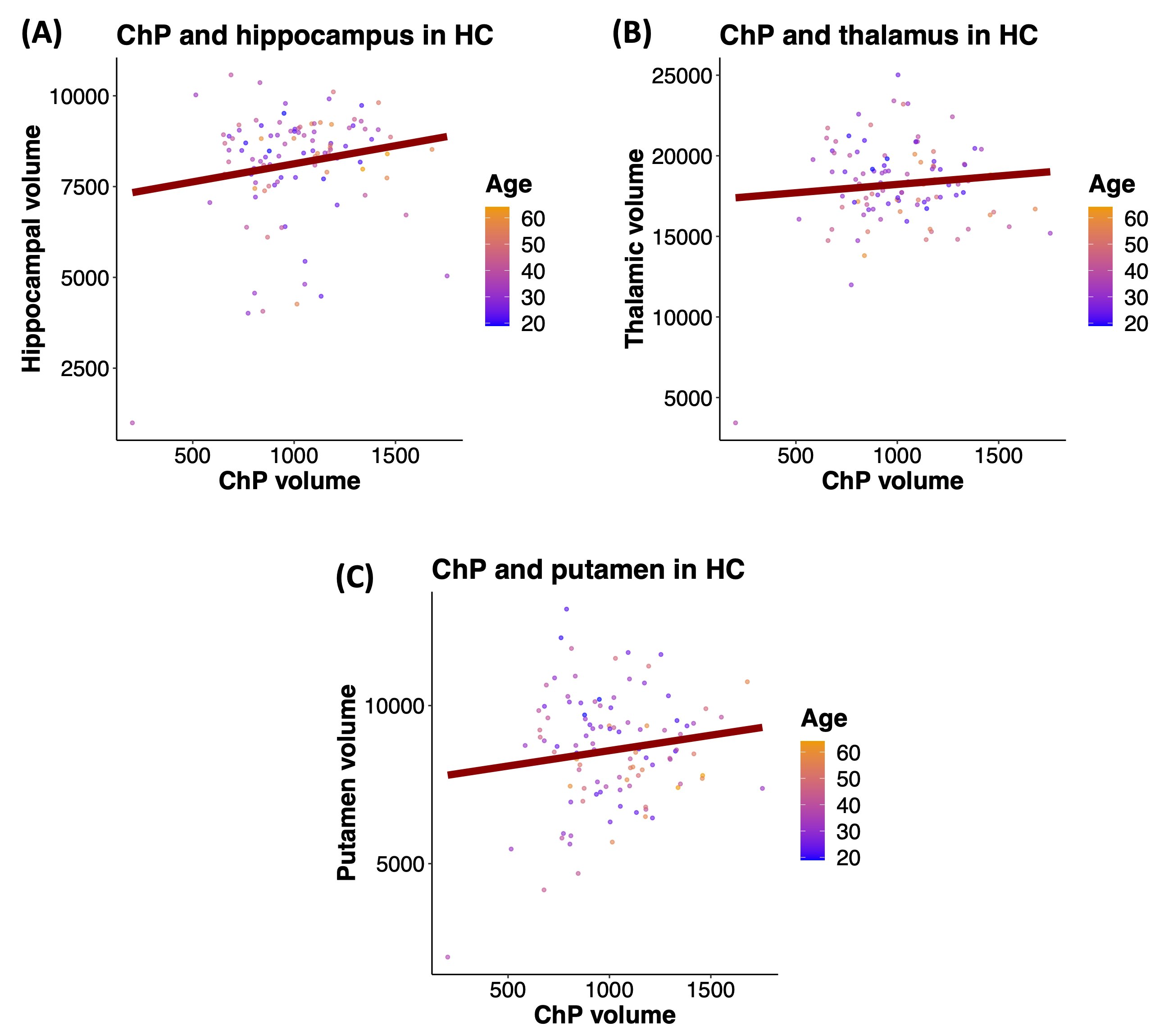

**Figure S7**

### Supplemental Tables

| **List of Abbreviations** | |
| --- | --- |
| **Abbreviation** | **Explanation** |
| AP | antipsychotic medication |
| BACS | Brief Assessment of Cognition in Schizophrenia |
| BMI | body mass index |
| BriefPD | brief psychotic disorder |
| ChP | choroid plexus |
| CI | confidence interval |
| DD | delusional disorder |
| DUP | duration of untreated psychosis |
| DUI | duration of illness |
| HC | healthy controls |
| hsCRP | high-sensitive C-reactive protein |
| LatVent | lateral ventricle |
| LL | lower limit |
| *N* | number of participants |
| *n* | number of plexi |
| *p* | *p* value |
| PANSS | Positive and Negative Syndrome Scale |
| PRS | polygenic risk score |
| *q* | false discovery rate adjusted *p* value |
| SE | standard error of mean |
| *SD* | standard deviation |
| SSD | schizophrenia spectrum disorder |
| SZ | schizophrenia |
| SZA | schizoaffective disorder |
| TRS | treatment resistant schizophrenia |
| UL | upper limit |
| VR | variability ratio |

**Table S1a. Descriptive statistics for choroid plexus measures.**

| **Region of interest** | **Mean, SSD** | ***SD*, SSD** | **N, SSD** | **Mean, HC** | ***SD*, HC** | **N, HC** |
| --- | --- | --- | --- | --- | --- | --- |
| Left choroid plexus (mm³) | 495.33 | 174.99 | 132 | 475.25 | 147.86 | 107 |
| Right choroid plexus (mm³) | 556.05 | 181.6 | 132 | 541.65 | 138.48 | 107 |

**Table S1b. Complete parameter estimates – group comparison of choroid plexus volume by hemisphere.**

| **Response** | **Predictor** | **Estimate** | **95% CI [LL, UL]** | ***N*, SSD** | ***n* (plexi), SSD** | ***N*, HC** | ***n* (plexi), HC** | ***p*** |
| --- | --- | --- | --- | --- | --- | --- | --- | --- |
| Choroid plexus (mm^3^) | (Intercept) | 360.38 | [290.9, 429.85] | 132 | 264 | 107 | 214 | **<0.001** |
| Choroid plexus (mm^3^) | Group (SSD vs HC) | 19.14 | [-15.37, 53.64] | 132 | 264 | 107 | 214 | 0.362 |
| Choroid plexus (mm^3^) | Age | 3.08 | [1.67. 4.5] | 132 | 264 | 107 | 214 | **<0.001** |
| Choroid plexus (mm^3^) | Sex (male vs female) | 0.91 | [-35.38, 37.21] | 132 | 264 | 107 | 214 | 0.967 |
| Choroid plexus (mm^3^) | Hemisphere (right vs left) | 66.41 | [43.72, 89.1] | 132 | 264 | 107 | 214 | **<0.001** |
| Choroid plexus (mm^3^) | Group x Hemisphere | -5.7 | [-36.22, 24.83] | 132 | 264 | 107 | 214 | 0.759 |

**Table S1c. Complete parameter estimates – group comparison of choroid plexus asymmetry index.**

| **Response** | **Predictor** | **Estimate** | **95% CI [LL, UL]** | ***N*, SSD** | ***N*, HC** | ***p*** |
| --- | --- | --- | --- | --- | --- | --- |
| ChP asymmetry index | (Intercept) | -0.088 | [-0.229, 0.054] | 132 | 107 | 0.308 |
| ChP asymmetry index | Group (SSD vs HC) | 0.027 | [-0.037, 0.091] | 132 | 107 | 0.486 |
| ChP asymmetry index | Age | -0.001 | [-0.04. 0.02] | 132 | 107 | 0.657 |
| ChP asymmetry index | Sex (male vs female) | -0.036 | [-0.111, 0.038] | 132 | 107 | 0.423 |

**Table S1d. Complete parameter estimates – association analysis of ChP volume with duration of antipsychotic treatment in SSD individuals.**

| **Response** | **Predictor** | **Estimate** | **95% CI [LL, UL]** | ***N*, SSD** | ***p*** |
| --- | --- | --- | --- | --- | --- |
| Left choroid plexus (mm^3^) | (Intercept) | 394.22 | [280.38, 508.07] | 121 | **<0.001** |
| Left choroid plexus (mm^3^) | AP treatment (months) | 0.14 | [-0.16, 0.44] | 121 | 0.433 |
| Left choroid plexus (mm^3^) | Sex (male vs female) | -18.62 | [-79.67, 42.43] | 121 | 0.614 |
| Left choroid plexus (mm^3^) | Age (years) | 2.78 | [-0.07, 5.63] | 121 | 0.109 |
| Right choroid plexus (mm^3^) | (Intercept) | 413.06 | [297.87, 528.25] | 121 | **<0.001** |
| Right choroid plexus (mm^3^) | AP treatment (months) | 0.21 | [-0.09, 0.51] | 121 | 0.238 |
| Right choroid plexus (mm^3^) | Sex (male vs female) | 4.38 | [-57.39, 66.15] | 121 | 0.907 |
| Right choroid plexus (mm^3^) | Age (years) | 3.03 | [0.15, 5.92] | 121 | 0.084 |

**Table S2a. Variability ratios of regional volumes and cortical thickness (bilateral).**

| **Regions of interest** | **VR (volume)** | **95% CI LL** | **95% CI UL** | **VR (thickness)** | **95% CI LL** | **95% CI UL** | ***N*, SSD** | ***N*, HC** |
| --- | --- | --- | --- | --- | --- | --- | --- | --- |
| Choroid plexus | 1.249 | 1.01 | 1.55 | - | - | - | 132 | 107 |
| Lateral ventricle | 1.364 | 1.1 | 1.69 | - | - | - | 132 | 107 |
| Third ventricle | 1.098 | 0.88 | 1.36 | - | - | - | 132 | 107 |
| Fourth ventricle | 0.823 | 0.66 | 1.02 | - | - | - | 132 | 107 |
| *Hippocampus* | 0.939 | 0.76 | 1.16 | - | - | - | 132 | 107 |
| *Anterior cingulate cortex* | 1.089 | 0.88 | 1.35 | 0.928 | 0.75 | 1.15 | 132 | 107 |
| *Posterior cingulate cortex* | 1.238 | 1 | 1.54 | 0.974 | 0.79 | 1.21 | 132 | 107 |
| *Insula* | 0.897 | 0.72 | 1.11 | 0.885 | 0.71 | 1.1 | 132 | 107 |
| *Cerebellum cortex* | 1.013 | 0.82 | 1.26 | - | - | - | 132 | 107 |
| *Fusiform gyrus* | 1.186 | 0.96 | 1.47 | 0.874 | 0.7 | 1.08 | 132 | 107 |
| *Superior temporal gyrus* | 1.084 | 0.87 | 1.34 | 0.94 | 0.76 | 1.17 | 132 | 107 |
| *Medial orbitofrontal cortex* | 1.071 | 0.86 | 1.33 | 0.833 | 0.67 | 1.03 | 132 | 107 |
| *Inferior temporal gyrus* | 1.005 | 0.81 | 1.25 | 0.966 | 0.78 | 1.2 | 132 | 107 |
| *Posterior parietal cortex* | 1.003 | 0.81 | 1.24 | 0.949 | 0.76 | 1.18 | 132 | 107 |
| *Lateral occipital cortex* | 1.028 | 0.83 | 1.28 | 1.005 | 0.81 | 1.25 | 132 | 107 |
| *Brainstem* | 0.788 | 0.64 | 0.98 | - | - | - | 132 | 107 |

**Table S2b. Variability ratios of regional volumes and cortical thickness (unilateral).**

| **Regions of interest** | **VR (volume)** | **95% CI LL** | **95% CI UL** | **VR (thickness)** | **95% CI LL** | **95% CI UL** | ***N*, SSD** | ***N*, HC** |
| --- | --- | --- | --- | --- | --- | --- | --- | --- |
| Left choroid plexus | 1.182 | 0.95 | 1.47 | - | - | - | 132 | 107 |
| Right choroid plexus | 1.31 | 1.06 | 1.63 | - | - | - | 132 | 107 |
| Left lateral ventricle | 1.469 | 1.18 | 1.82 | - | - | - | 132 | 107 |
| Right lateral ventricle | 1.199 | 0.97 | 1.49 | - | - | - | 132 | 107 |
| *Left hippocampus* | 0.951 | 0.77 | 1.18 | - | - | - | 132 | 107 |
| *Right hippocampus* | 0.862 | 0.71 | 1.05 | - | - | - | 132 | 107 |
| *Left caudal anterior cingulate* | 1.271 | 1.02 | 1.58 | 0.999 | 0.81 | 1.24 | 132 | 132 |
| *Right caudal anterior cingulate* | 1.104 | 0.89 | 1.37 | 0.982 | 0.79 | 1.22 | 132 | 132 |
| *Left rostral anterior cingulate* | 0.909 | 0.73 | 1.13 | 0.827 | 0.67 | 1.03 | 132 | 132 |
| *Right rostral anterior cingulate* | 1.189 | 0.96 | 1.47 | 0.971 | 0.78 | 1.2 | 132 | 132 |
| *Left posterior cingulate* | 1.171 | 0.94 | 1.45 | 0.969 | 0.78 | 1.2 | 132 | 107 |
| *Right posterior cingulate* | 1.248 | 1.01 | 1.55 | 0.987 | 0.8 | 1.23 | 132 | 107 |
| *Left insula* | 0.749 | 0.6 | 0.93 | 0.849 | 0.68 | 1.05 | 132 | 107 |
| *Right insula* | 1.036 | 0.83 | 1.29 | 0.936 | 0.75 | 1.16 | 132 | 107 |
| *Left cerebellum cortex* | 1.045 | 0.84 | 1.3 | - | - | - | 132 | 107 |
| *Right cerebellum cortex* | 0.884 | 0.71 | 1.1 | - | - | - | 132 | 107 |
| *Left fusiform gyrus* | 1.208 | 0.97 | 1.5 | 0.864 | 0.7 | 1.07 | 132 | 107 |
| *Right fusiform gyrus* | 1.11 | 0.89 | 1.38 | 0.901 | 0.73 | 1.12 | 132 | 107 |
| *Left superior temporal gyrus* | 1.076 | 0.87 | 1.34 | 0.957 | 0.77 | 1.19 | 132 | 107 |
| *Right superior temporal gyrus* | 1.113 | 0.9 | 1.38 | 0.931 | 0.75 | 1.16 | 132 | 107 |
| *Left medial orbitofrontal cortex* | 0.902 | 0.73 | 1.12 | 0.829 | 0.67 | 1.03 | 132 | 107 |
| *Right medial orbitofrontal cortex* | 1.245 | 1 | 1.55 | 0.836 | 0.67 | 1.04 | 132 | 107 |
| *Left inferior temporal gyrus* | 0.907 | 0.73 | 1.12 | 0.98 | 0.79 | 1.22 | 132 | 107 |
| *Right inferior temporal gyrus* | 1.149 | 0.93 | 1.43 | 0.961 | 0.77 | 1.19 | 132 | 107 |
| *Left superior parietal cx* | 1.075 | 0.87 | 1.33 | 0.97 | 0.78 | 1.2 | 132 | 107 |
| *Right superior parietal cx* | 1.099 | 0.89 | 1.36 | 0.948 | 0.76 | 1.18 | 132 | 107 |
| *Left inferior parietal cx* | 0.86 | 0.69 | 1.07 | 0.999 | 0.81 | 1.24 | 132 | 132 |
| *Right inferior parietal cx* | 0.901 | 0.73 | 1.12 | 0.889 | 0.72 | 1.1 | 132 | 132 |
| *Left lateral occipital cortex* | 0.966 | 0.78 | 1.2 | 0.99 | 0.8 | 1.23 | 132 | 107 |
| *Right lateral occipital cortex* | 1.081 | 0.87 | 1.34 | 1.009 | 0.81 | 1.25 | 132 | 107 |

**Table S3a. General demographic and clinical cluster characteristics.**

| **Cluster** | **Cluster description** | **Scaled cluster central points (ChP/LatVent)** | **N** | **Sex (f/m)** | **Diagnosis (SZ/SZA/BriefPD/DD)** | **Treatment resistance (TRS/non-TRS)** | **Smoking status (non-smokers/smokers)** |
| --- | --- | --- | --- | --- | --- | --- | --- |
| 1 | "large ChP-large ventricle cluster" | 1.314/2.366 | 11 | 1/10 | 6/5/0/0 | 5/4 | 5/6 |
| 2 | "small ChP-small ventricle cluster" | -0.76/-0.549 | 65 | 14/51 | 49/11/5/0 | 19/45 | 26/35 |
| 3 | "intermediate ChP-small ventricle cluster" | 0.624/0.173 | 56 | 16/40 | 40/14/1/1 | 23/31 | 32/23 |

**Table S3b. General demographic and clinical cluster characteristics.**

|  | **Cluster 1** | | **Cluster 2** | | **Cluster 3** | |  |  |  |
| --- | --- | --- | --- | --- | --- | --- | --- | --- | --- |
| **Variable** | **Mean** | **SD** | **Mean** | **SD** | **Mean** | **SD** | **p** | **q** | **GH post-hoc** |
| GAF score | 46.5 | 11.9 | 51.6 | 13.1 | 55.8 | 8.89 | **0.022** | 0.055 |  |
| Age (years) | 47.7 | 9.62 | 32.9 | 9.55 | 40.4 | 11 | **<0.001** | **<0.001** | 1>2***, 3>2***, 1>3. |
| BMI (kg/m^2^) | 28.3 | 6.8 | 27.5 | 5.2 | 28.7 | 4.63 | 0.426 | 0.426 |  |
| DUI (months) | 197 | 130 | 123 | 112 | 156 | 113 | 0.088 | 0.147 |  |
| DUP (months) | 24.3 | 21.6 | 27.3 | 42 | 14.2 | 24.1 | 0.292 | 0.365 |  |

**Table S3c. Psychopathology-related cluster characteristics.**

|  | **Cluster 1** | | **Cluster 2** | | **Cluster 3** | |  |  |
| --- | --- | --- | --- | --- | --- | --- | --- | --- |
| **Variable** | **Mean** | **SD** | **Mean** | **SD** | **Mean** | **SD** | **p** | **q** |
| PANSS total score | 60.1 | 17.1 | 55.8 | 14.2 | 52.6 | 15.8 | 0.234 | 0.312 |
| PANSS positive score | 12.8 | 4.87 | 12.9 | 4.05 | 12.7 | 5.15 | 0.986 | 0.986 |
| PANSS negative score | 15.5 | 5.13 | 14 | 5.75 | 12.5 | 4.91 | 0.113 | 0.312 |
| PANSS general score | 31.7 | 8.74 | 28.9 | 7.37 | 27.4 | 7.97 | 0.185 | 0.312 |

**Table S3d. Cognition-related cluster characteristics.**

|  | **Cluster 1** | | **Cluster 2** | | **Cluster 3** | |  |  |  |
| --- | --- | --- | --- | --- | --- | --- | --- | --- | --- |
| **Variable** | **Mean** | **SD** | **Mean** | **SD** | **Mean** | **SD** | **p** | **q** | **GH post-hoc** |
| BACS total composite z score | -1.48 | 1.39 | -0.52 | 0.735 | -0.558 | 1.12 | **0.033** | 0.077 |  |
| BACS digit sequence z score | -1.76 | 1.14 | -0.833 | 1.08 | -0.937 | 1.23 | 0.091 | 0.128 |  |
| BACS symbol coding z score | -2.38 | 1.11 | -1.15 | 0.923 | -1.26 | 1.19 | **0.007** | **0.025** | 2>1*, 3>1* |
| BACS token motor z score | -1.81 | 1.11 | -1.01 | 0.909 | -1.12 | 1.07 | 0.092 | 0.129 |  |
| BACS tower of London z score | -3 | 3.34 | -0.405 | 1.05 | -0.599 | 1.72 | **<0.001** | **0.001** |  |
| BACS verbal fluency z score | -0.804 | 1.21 | -0.798 | 0.913 | -0.609 | 1.14 | 0.624 | 0.624 |  |
| BACS verbal memory z score | -2.03 | 1.98 | -1.17 | 1.22 | -1.29 | 1.62 | 0.268 | 0.313 |  |

**Table S3e. Inflammation-related cluster characteristics.**

|  | **Cluster 1** | | **Cluster 2** | | **Cluster 3** | |  |  |  |
| --- | --- | --- | --- | --- | --- | --- | --- | --- | --- |
| **Variable** | **Mean** | **SD** | **Mean** | **SD** | **Mean** | **SD** | **p** | **q** | **GH post-hoc** |
| Neutrophils (G/l) | 4.93 | 1.18 | 3.72 | 1.2 | 4.44 | 1.33 | **0.016** | **0.044** | 3>2., 1>2. |
| Monocytes (G/l) | 0.637 | 0.14 | 0.517 | 0.145 | 0.496 | 0.12 | **0.033** | **0.044** |  |
| Lymphocytes (G/l) | 2.27 | 0.58 | 1.85 | 0.61 | 1.93 | 0.652 | 0.321 | 0.321 |  |
| hsCRP (pg/ml) | 3.78×10^8^ | 3.94×10^8^ | 2.20×10^8^ | 2.35×10^8^ | 1.60×10^8^ | 1.49×10^8^ | **0.026** | **0.044** |  |

**Table S3f. Between-cluster differences in genetic liability for schizophrenia.**

|  | **Cluster 1** | | **Cluster 2** | | **Cluster 3** | |  |
| --- | --- | --- | --- | --- | --- | --- | --- |
| **Variable** | **Mean** | **SD** | **Mean** | **SD** | **Mean** | **SD** | **p** |
| SZ-PRS | -6.52 | 0.173 | -6.47 | 0.246 | -6.43 | 0.261 | 0.562 |

**Table S4a. Complete parameter estimates – association analysis of ChP with subcortical regions in SSD participants.**

| **Response** | **Predictor** | **Estimate** | **95% CI [LL, UL]** | ***N*, SSD** | ***p*** | ***q*** |
| --- | --- | --- | --- | --- | --- | --- |
| Hippocampus (mm^3^) | (Intercept) | 7580.99 | [6658.91, 8503.07] | 132 | **<0.001** | **<0.001** |
| Hippocampus (mm^3^) | Choroid plexus (mm^3^) | 1.41 | [0.78, 2.03] | 132 | **<0.001** | **0.001** |
| Hippocampus (mm^3^) | Sex (male vs female) | -601.46 | [-1055.25, -147.67] | 132 | **0.03** | **0.04** |
| Hippocampus (mm^3^) | Age (years) | -24 | [-42, -6] | 132 | **0.029** | 0.058 |
| Amygdala (mm^3^) | (Intercept) | 3085.73 | [2723.64, 3447.83] | 132 | **<0.001** | **<0.001** |
| Amygdala (mm^3^) | Choroid plexus (mm^3^) | 0.12 | [-0.13, 0.36] | 132 | 0.438 | 0.438 |
| Amygdala (mm^3^) | Sex (male vs female) | -240.55 | [-418.75, -62.35] | 132 | **0.027** | **0.04** |
| Amygdala (mm^3^) | Age (years) | -4.36 | [-11.43, 2.71] | 132 | 0.309 | 0.412 |
| Putamen (mm^3^) | (Intercept) | 10303.95 | [9104.23, 11503.67] | 132 | **<0.001** | **<0.001** |
| Putamen (mm^3^) | Choroid plexus (mm^3^) | 1.18 | [0.37, 2] | 132 | **0.018** | **0.024** |
| Putamen (mm^3^) | Sex (male vs female) | -835.26 | [-1425.68, -244.83] | 132 | **0.021** | **0.04** |
| Putamen (mm^3^) | Age (years) | -60.15 | [-83.57, 36.74] | 132 | **<0.001** | **<0.001** |
| Thalamus (mm^3^) | (Intercept) | 15815.78 | [13953.71, 17677.84] | 132 | **<0.001** | **<0.001** |
| Thalamus (mm^3^) | Choroid plexus (mm^3^) | 1.932 | [0.67, 3.2] | 132 | **0.013** | **0.024** |
| Thalamus (mm^3^) | Sex (male vs female) | 438.09 | [-478.3, 1354.48] | 132 | 0.43 | 0.43 |
| Thalamus (mm^3^) | Age (years) | -17.79 | [-54.13, 18.56] | 132 | 0.419 | 0.419 |

**Table S4b. Complete parameter estimates – association analysis of ChP with regional cortical volumes in SSD participants.**

| **Response** | **Predictor** | **Estimate** | **95% CI [LL, UL]** | ***N*, SSD** | ***p*** | ***q*** |
| --- | --- | --- | --- | --- | --- | --- |
| Superior temporal gyrus (mm^3^) | (Intercept) | 34219.58 | [31161.14, 37278.03] | 132 | **<0.001** | **<0.001** |
| Superior temporal gyrus (mm^3^) | Choroid plexus (mm^3^) | 0.165 | [-1.911, 2.241] | 132 | 0.895 | 0.935 |
| Superior temporal gyrus (mm^3^) | Sex (male vs female) | -353.13 | [-1858.31, 1152.05] | 132 | 0.698 | 0.698 |
| Superior temporal gyrus (mm^3^) | Age (years) | -87.04 | [-146.74, -27.34] | 132 | **0.017** | **0.028** |
| Superior frontal gyrus (mm^3^) | (Intercept) | 57109.29 | [51674.2, 62544.38] | 132 | **<0.001** | **<0.001** |
| Superior frontal gyrus (mm^3^) | Choroid plexus (mm^3^) | 1.51 | [-1.65, 1.82] | 132 | 0.498 | 0.935 |
| Superior frontal gyrus (mm^3^) | Sex (male vs female) | 1288.05 | [-1386.76, 3962.87] | 132 | 0.426 | 0.698 |
| Superior frontal gyrus (mm^3^) | Age (years) | -242.15 | [-348.24, -136.06] | 132 | **<0.001** | **<0.001** |
| Precentral gyrus (mm^3^) | (Intercept) | 28648.55 | [26087.31, 31209.78] | 132 | **<0.001** | **<0.001** |
| Precentral gyrus (mm^3^) | Choroid plexus (mm^3^) | 0.086 | [0.37, 2] | 132 | 0.935 | 0.935 |
| Precentral gyrus (mm^3^) | Sex (male vs female) | 501.4 | [-759.08,1761.88] | 132 | 0.511 | 0.698 |
| Precentral gyrus (mm^3^) | Age (years) | -69.79 | [-119.79, -19.8] | 132 | **0.022** | **0.028** |
| Fusiform gyrus (mm^3^) | (Intercept) | 18860.46 | [17068.3, 20652.62] | 132 | **<0.001** | **<0.001** |
| Fusiform gyrus (mm^3^) | Choroid plexus (mm^3^) | 0.271 | [-0.95, 1.49] | 132 | 0.712 | 0.935 |
| Fusiform gyrus (mm^3^) | Sex (male vs female) | -217.29 | [-1099.28, 664.7] | 132 | 0.684 | 0.698 |
| Fusiform gyrus (mm^3^) | Age (years) | -117.87 | [-152.86, -82.89] | 132 | **<0.001** | **<0.001** |
| Caudal middle frontal gyrus (mm^3^) | (Intercept) | 14693.4 | [12672.72, 16714.09] | 132 | **<0.001** | **<0.001** |
| Caudal middle frontal gyrus (mm^3^) | Choroid plexus (mm^3^) | 0.31 | [-1.06, 1.68] | 132 | 0.707 | 0.935 |
| Caudal middle frontal gyrus (mm^3^) | Sex (male vs female) | 499.29 | [-495.17, 1493.74] | 132 | 0.407 | 0.698 |
| Caudal middle frontal gyrus (mm^3^) | Age (years) | -40.05 | [-79.49, -0.61] | 132 | 0.095 | 0.095 |

**Table S4c. Complete parameter estimates – association analysis of lateral ventricle with subcortical regions in SSD participants.**

| **Response** | **Predictor** | **Estimate** | **95% CI [LL, UL]** | ***N*, SSD** | ***p*** | ***q*** |
| --- | --- | --- | --- | --- | --- | --- |
| Hippocampus (mm^3^) | (Intercept) | 8622.67 | [7786.49, 9458.85] | 132 | **<0.001** | **<0.001** |
| Hippocampus (mm^3^) | Lateral ventricle (mm^3^) | -0.009 | [-0.032, 0.013] | 132 | 0.486 | 0.489 |
| Hippocampus (mm^3^) | Sex (male vs female) | -566.95 | [-1053.4, --80.5] | 132 | 0.056 | 0.084 |
| Hippocampus (mm^3^) | Age (years) | -8.39 | [-29.12, 12.34] | 132 | 0.504 | 0.673 |
| Putamen (mm^3^) | (Intercept) | 11176.94 | [10122.99, 12230.88] | 132 | **<0.001** | **<0.001** |
| Putamen (mm^3^) | Lateral ventricle (mm^3^) | -0.018 | [-0.046, 0.01] | 132 | 0.285 | 0.489 |
| Putamen (mm^3^) | Sex (male vs female) | -761.59 | [-1374.72, -148.45] | 132 | **0.042** | 0.084 |
| Putamen (mm^3^) | Age (years) | -42.39 | [-68.51, -16.26] | 132 | **0.008** | **0.024** |
| Thalamus (mm^3^) | (Intercept) | 17260.15 | [15616.25, 18904.04] | 132 | **<0.001** | **<0.001** |
| Thalamus (mm^3^) | Lateral ventricle (mm^3^) | 0.018 | [-0.026, 0.062] | 132 | 0.489 | 0.489 |
| Thalamus (mm^3^) | Sex (male vs female) | 349.91 | [-606.43, 1306.24] | 132 | 0.545 | 0.545 |
| Thalamus (mm^3^) | Age (years) | -10.39 | [-51.14, 30.36] | 132 | 0.673 | 0.673 |

**Table S5. Complete parameter estimates – association analysis of ChP with subcortical regions in HC participants.**

| **Response** | **Predictor** | **Estimate** | **95% CI [LL, UL]** | ***N*, SSD** | ***p*** | ***q*** |
| --- | --- | --- | --- | --- | --- | --- |
| Hippocampus (mm^3^) | (Intercept) | 8693.11 | [7279.53, 10106.69] | 107 | **<0.001** | **<0.001** |
| Hippocampus (mm^3^) | Choroid plexus (mm^3^) | 0.919 | [0, 1.84] | 107 | 0.1 | 0.15 |
| Hippocampus (mm^3^) | Sex (male vs female) | -1070.97 | [-1652.76, -489.17] | 107 | **0.003** | **0.009** |
| Hippocampus (mm^3^) | Age (years) | -20.86 | [-44.37, 2.65] | 107 | 0.144 | 0.216 |
| Putamen (mm^3^) | (Intercept) | 10198.84 | [9104.23, 11503.67] | 107 | **<0.001** | **<0.001** |
| Putamen (mm^3^) | Choroid plexus (mm^3^) | 1.2 | [0.13, 2.26] | 107 | 0.064 | 0.15 |
| Putamen (mm^3^) | Sex (male vs female) | -1072.52 | [-1745.22, -399.81] | 107 | **0.009** | **0.014** |
| Putamen (mm^3^) | Age (years) | -57 | [-84.18, -29.82] | 107 | **0.001** | **0.003** |
| Thalamus (mm^3^) | (Intercept) | 16624.06 | [14064.17, 19183.96] | 107 | **<0.001** | **<0.001** |
| Thalamus (mm^3^) | Choroid plexus (mm^3^) | 1.39 | [-0.27, 3.05] | 107 | 0.169 | 0.169 |
| Thalamus (mm^3^) | Sex (male vs female) | 1041.73 | [-11.86, 2095.33] | 107 | 0.104 | 0.104 |
| Thalamus (mm^3^) | Age (years) | -13.14 | [-55.72, 29.44] | 107 | 0.61 | 0.61 |

### Supplemental References

**1.**	Roell L, Keeser D, Papazov B, et al. Effects of Exercise on Structural and Functional Brain Patterns in Schizophrenia-Data From a Multicenter Randomized-Controlled Study. *Schizophr Bull* Aug 19 2023.

**2.**	Esteban O, Birman D, Schaer M, Koyejo OO, Poldrack RA, Gorgolewski KJ. MRIQC: Advancing the automatic prediction of image quality in MRI from unseen sites. *PLoS One* 2017;12(9):e0184661.

**3.**	Reuter M, Rosas HD, Fischl B. Highly accurate inverse consistent registration: a robust approach. *Neuroimage* Dec 2010;53(4):1181-1196.

**4.**	Segonne F, Dale AM, Busa E, Glessner M, Salat D, Hahn HK, Fischl B. A hybrid approach to the skull stripping problem in MRI. *Neuroimage* Jul 2004;22(3):1060-1075.

**5.**	Fischl B, Salat DH, Busa E, et al. Whole brain segmentation: automated labeling of neuroanatomical structures in the human brain. *Neuron* Jan 31 2002;33(3):341-355.

**6.**	Fischl B, Salat DH, van der Kouwe AJ, Makris N, Segonne F, Quinn BT, Dale AM. Sequence-independent segmentation of magnetic resonance images. *Neuroimage* 2004;23 Suppl 1:S69-84.

**7.**	Sled JG, Zijdenbos AP, Evans AC. A nonparametric method for automatic correction of intensity nonuniformity in MRI data. *IEEE Trans Med Imaging* Feb 1998;17(1):87-97.

**8.**	Fischl B, Liu A, Dale AM. Automated manifold surgery: constructing geometrically accurate and topologically correct models of the human cerebral cortex. *IEEE Trans Med Imaging* Jan 2001;20(1):70-80.

**9.**	Segonne F, Pacheco J, Fischl B. Geometrically accurate topology-correction of cortical surfaces using nonseparating loops. *IEEE Trans Med Imaging* Apr 2007;26(4):518-529.

**10.**	Dale AM, Fischl B, Sereno MI. Cortical surface-based analysis. I. Segmentation and surface reconstruction. *Neuroimage* Feb 1999;9(2):179-194.

**11.**	Dale AM, Sereno MI. Improved Localizadon of Cortical Activity by Combining EEG and MEG with MRI Cortical Surface Reconstruction: A Linear Approach. *J Cogn Neurosci* Spring 1993;5(2):162-176.

**12.**	Fischl B, Dale AM. Measuring the thickness of the human cerebral cortex from magnetic resonance images. *Proc Natl Acad Sci U S A* Sep 26 2000;97(20):11050-11055.

**13.**	Jernigan TL, Zatz LM, Moses JA, Jr., Berger PA. Computed tomography in schizophrenics and normal volunteers. I. Fluid volume. *Arch Gen Psychiatry* Jul 1982;39(7):765-770.

**14.**	Papiol S, Keeser D, Hasan A, et al. Polygenic burden associated to oligodendrocyte precursor cells and radial glia influences the hippocampal volume changes induced by aerobic exercise in schizophrenia patients. *Transl Psychiatry* Nov 11 2019;9(1):284.

**15.**	Das S, Forer L, Schonherr S, et al. Next-generation genotype imputation service and methods. *Nat Genet* Oct 2016;48(10):1284-1287.

**16.**	Trubetskoy V, Pardiñas AF, Qi T, et al. Mapping genomic loci implicates genes and synaptic biology in schizophrenia. *Nature* Apr 2022;604(7906):502-508.

**17.**	Ge T, Chen CY, Ni Y, Feng YA, Smoller JW. Polygenic prediction via Bayesian regression and continuous shrinkage priors. *Nat Commun* Apr 16 2019;10(1):1776.

**18.**	Boudriot E, Gabriel V, Popovic D, et al. Signature of altered retinal microstructures and electrophysiology in schizophrenia spectrum disorders is associated with disease severity and polygenic risk. *Biol Psychiatry* Apr 26 2024.

**19.**	Schijven D, Postema MC, Fukunaga M, et al. Large-scale analysis of structural brain asymmetries in schizophrenia via the ENIGMA consortium. *Proc Natl Acad Sci U S A* Apr 4 2023;120(14):e2213880120.

**20.**	Kong XZ, Mathias SR, Guadalupe T, et al. Mapping cortical brain asymmetry in 17,141 healthy individuals worldwide via the ENIGMA Consortium. *Proc Natl Acad Sci U S A* May 29 2018;115(22):E5154-e5163.
